# Utilizing nullomers in cell-free RNA for early cancer detection

**DOI:** 10.1101/2023.06.10.23291228

**Authors:** Austin Montgomery, Georgios Christos Tsiatsianis, Ioannis Mouratidis, Candace S.Y. Chan, Maria Athanasiou, Anastasios D. Papanastasiou, Verena Kantere, Ioannis Vathiotis, Konstantinos Syrigos, Nelson S. Yee, Ilias Georgakopoulos-Soares

## Abstract

Early detection of cancer can significantly improve patient outcomes; however, sensitive and highly specific biomarkers for cancer detection are currently missing. Nullomers are short sequences that are absent from the human genome but can resurface due to somatic mutations in cancer. We examine over 10,000 whole exome sequencing matched tumor-normal samples to characterize nullomer resurfacing across exonic regions of the genome. We identify nullomer resurfacing mutational hotspots within tumor genes and report that certain mutational signatures are associated with nullomer resurfacing. We show that DNA mismatch repair and homologous recombination repair can be detected from the nullomer profile and provide evidence that nullomers can be used to identify neoepitopes and other targets for precision oncology. Finally, we provide evidence for the identification of nullomers in cell free RNA from peripheral blood samples, enabling early detection of multiple tumor types. We show multiple tumor classification models with an AUC greater than 0.9, including a hepatocellular carcinoma classifier with an AUC greater than 0.99.

## Introduction

Cancer is characterized by the accumulation of somatic mutations and uncontrolled clonal proliferation of malignant cells (Stratton, Campbell, and Futreal 2009). Even though there have been important advances in cancer therapeutics, cancer remains the second leading cause of death worldwide (Siegel, Miller, and Jemal 2020). Early cancer detection is associated with improved clinical outcomes (Crosby et al. 2022). Nevertheless, in the vast majority of cases, malignant tumors are detected at a late stage, where the likelihood of survival declines steeply (Mattiuzzi and Lippi 2019). As a result, there is an unmet need for the development of novel biomarkers, which will enable early cancer detection as well as surveillance at the population level.

Cancer biomarker development has involved proteomic, transcriptomic and metabolomic profiling, DNA methylation, circulating tumor cells, and cell-free DNA (cfDNA) (Rushton et al. 2021; Zaporozhchenko et al. 2018; Nielsen 2017; Locke et al. 2019; Ding et al. 2022). However, these methods have been shown to have suboptimal sensitivity and specificity. There is also sufficient evidence that cancer cells release cfRNA, which can be detected in the blood. There is also sufficient evidence that cancer cells release cfRNA, which can be detected in the blood (Larson et al. 2022). cfRNA represents a highly dynamic biomarker, since it can indicate expression changes in real time. Importantly, highly expressed tumor-associated genes can be over-represented in cfRNA samples relative to their lower frequency in cfDNA. cfRNA can also provide information about the tissue of origin as there are tissue-specific and cancer-specific transcriptomic differences (Larson et al. 2022). Consequently, cfRNA can provide information that is complementary to that derived from cfDNA and could prove particularly useful for tumors with lower mutational load.

Nullomers are sequences that are absent from the human genome (Acquisti et al. 2007). We and others have previously genomically characterized nullomers and provided evidence for negative selection constraints and for resurfacing due to germline variants (Georgakopoulos-Soares, Yizhar-Barnea, et al. 2021; Koulouras and Frith 2021). Recently, we have also investigated the relevance of nullomers in cancer; by analyzing more than 2,700 Whole Genome Sequenced primary tumors we provided evidence for the resurfacing of nullomers during cancer development while also showing the effectiveness of nullomers as early cancer detection biomarkers using cfDNA (Georgakopoulos-Soares, Barnea, et al. 2021). Even though exonic regions are enriched for nullomer resurfacing mutations, it is still unclear whether nullomers can be used for the early detection of cancer in cfRNA or carry prognostic and/or predictive relevance. The identification of tumor vulnerability targets, such as DNA mismatch repair deficiency (Pengfei Zhao et al. 2019), homologous recombination deficiency (Konstantinopoulos et al. 2015), or other actionable targets, can lead to personalized treatments with improved clinical outcomes (Lone et al. 2022). Thus, it remains a goal to be able to derive such information directly from liquid biopsies.

Along these lines, we were interested to examine nullomers’ utility as novel cfRNA biomarkers for early cancer detection, as well as a prognostic tool. Here, we perform an extensive analysis of nullomer resurfacing across more than 10,000 Whole Exome Sequencing (WES) tumor samples (Ellrott et al. 2018). We evaluate the distribution of nullomer resurfacing events across tumor types and patients and identify recurrent nullomer resurfacing events within cancer genes. We find that the mutational signature profile differs between mutations that cause nullomer resurfacing and mutations that do not cause nullomer resurfacing, and these findings hold across multiple cancer types. We also show that nullomers can be used for the detection of repair enzyme deficiencies and can thus serve as prognostic and predictive biomarkers. Finally, we use cfRNA data obtained from liquid biopsy samples to detect cancer using nullomers. Our findings provide evidence for the utility of nullomers as cancer diagnostic, prognostic and predictive biomarkers in cfRNA.

## Results

### Mutation type preferences during nullomer resurfacing in cancer

Even though nullomer sequences are absent from the human genome, somatic mutations can cause the resurfacing of nullomers during cancer development. We first identified nullomers across kmer lengths of up to 16 base-pairs (bp) long for the reference human genome as previously described in (Georgakopoulos-Soares, Barnea, et al. 2021). We analyzed mutation data from over 10,000 WES matched tumor-normal pairs across 32 cancer types, to detect resurfacing of nullomers due to somatic mutations. Germline mutations were not included in the analysis and were removed using the tumor-normal pairs. The total number of different sixteen bp nullomers that resurfaced across all somatic mutations in this cohort was 29,774,302, representing 0.69% of the sixteen bp kmer space. Moreover, we found that the proportion of somatic mutations that cause nullomer resurfacing increased from 0.178% at 12bp kmer length, to 79.76% at 16bp kmer length (**Figure 1a**). This finding indicates that at longer kmer lengths the majority of exonic somatic mutations cause the resurfacing of one or more nullomers.

**Figure 1:**
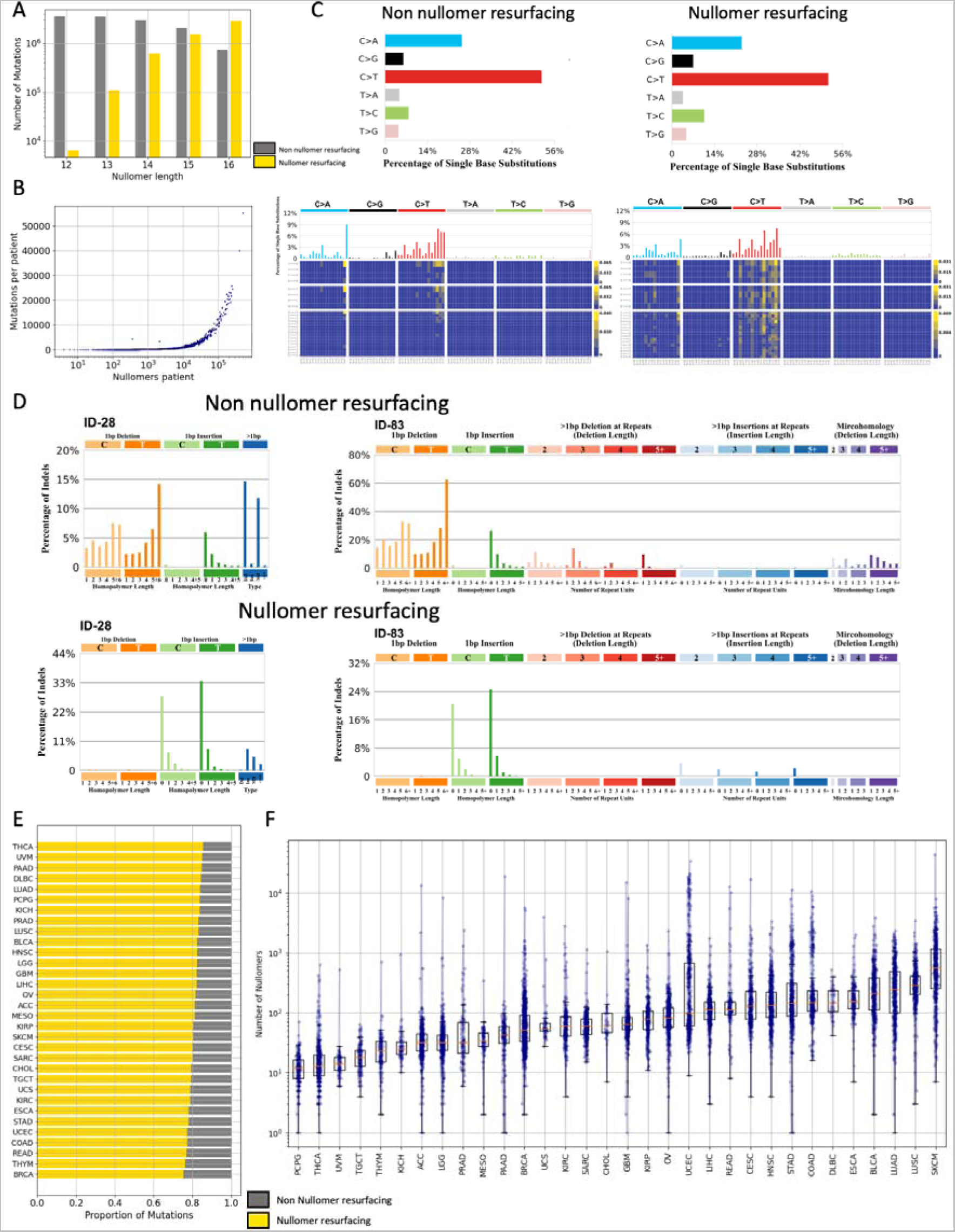
Characterization of nullomer resurfacing in WES patient samples. **A.** The number of mutations causing nullomer resurfacing (in yellow) relative to those that do not cause nullomer resurfacing for kmer lengths between 12bp and 16bp. Β. Association between the number of WES somatic mutations and the number of nullomers that resurface per patient. Results shown for sixteen-mer nullomers. **C.** Proportion of substitution types in nullomer resurfacing and non-nullomer resurfacing substitutions for sixteen bp nullomer length. Characterization of differences in substitution type preference for non-nullomer and nullomer resurfacing mutations using the 96 substitution type channels. **D.** Characterization of differences in indel preference for non-nullomer and nullomer resurfacing mutations using the 28 and 83 indel mutation channels. **E.** Proportion of mutations (in yellow) causing sixteen bp nullomer resurfacing across cancer types. The proportion of mutations that do not cause nullomer resurfacing are shown in gray. **F.** Number of nullomers detected for each cancer patient in each cancer type for 16bp nullomer length. Every dot represents a patient.

We also report a strong correlation between the number of mutations and the number of resurfaced nullomers across patients with cancer (Pearson correlation, r>0.98, p-value<0.0001 across kmer lengths; **Figure 1b**, **Supplementary Figure 1**). In addition, the average number of nullomers that resurface by each individual mutation increased with the nullomer length (**Supplementary Figure 2**). We also examined potential differences in the mutation types that are more likely to cause nullomer resurfacing. For substitutions, significant differences were revealed particularly when examining mutations using the 96 possible trinucleotide changes, stemming from the six substitution types (**Figure 1c**). For example, we observed that nullomer resurfacing mutations show a smaller proportion being TCT>TAT and a larger proportion being GCG>GTG (**Figure 1c**). We also explored indels and doublet base substitutions for nullomer resurfacing. We found that mononucleotide repeat tract deletions almost never cause nullomer resurfacing but instead nullomer resurfacing occurred primarily at 0bp or 1bp homopolymer length insertions (**Figure 1d**, **Supplementary Figure 3**). These findings indicate that the mutation type significantly influences the likelihood of nullomer resurfacing.

**Figure 2:**
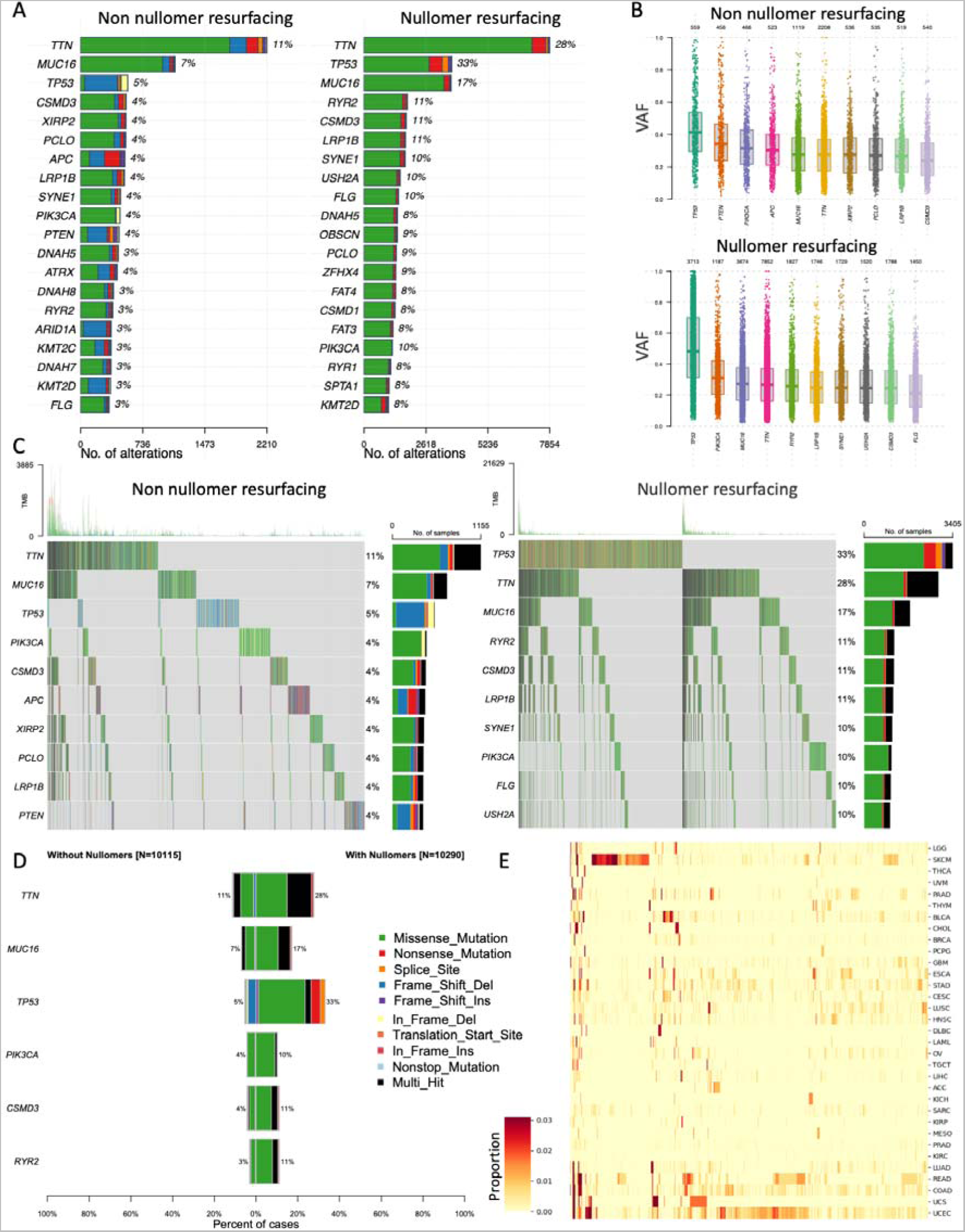
Identification of nullomer resurfacing across cancer genes. **A.** Percentage of patients with each mutated gene across cancer types for the top twenty most mutated genes. **B.** Variant allele frequency of mutations that do not cause sixteen-mer nullomer resurfacing and of mutations that cause sixteen-mer nullomer resurfacing. **C-D.** Number of mutations in top cancer genes for mutations that either do not cause or cause nullomer resurfacing. **E.** Proportion of patients in which each of the top 16bp top 10,000 nullomers from across all patients is found.

### Identification of nullomer resurfacing across 10,000 WES tumor samples

We investigated how the proportion of mutations that caused the resurfacing of nullomers changed as a function of kmer length at individual cancer types. The proportion of mutations that caused nullomer resurfacing was extremely small at twelve and thirteen bp lengths, whereas at sixteen bp lengths the majority of somatic mutations caused nullomer resurfacing across cancer types (**Figure 1e**, **Supplementary Figure 4**). We also report differences in the proportion of mutations causing nullomer resurfacing between cancer types, with thyroid cancer (THCA) having the highest and breast cancer (BRCA) the lowest proportion of mutations causing 16bp nullomer resurfacing (85.55% and 75.50% of mutations, respectively) and the findings were replicated across the other kmer lengths (**Figure 1e-f**, **Supplementary Figure 4**).

Next, the number of nullomers identified across individual cancer types and patients was explored. The mean number of nullomers identified across patients ranged between 0.62 and 278.5 for 12bp and 16bp kmer lengths respectively, and the cancer types with the highest and lowest number of nullomers resurfacing per patient were skin cutaneous melanoma (SKCM) and pheochromocytomas and paragangliomas (PCPG), respectively (**Figure 1f**). We also observed that in the most extreme case one patient produced 508,100 nullomers, indicating a hypermutator phenotype. We conclude that nullomer resurfacing occurred for a significant fraction of somatic mutations across cancer types, when examining kmer lengths of fourteen bps or higher.

### Nullomer resurfacing across cancer genes

Subsequently, we examined the frequency of nullomer resurfacing mutations and compared it to the frequency of mutations that did not cause nullomer resurfacing across genes. Firstly, across genes that were most frequently mutated in the patient cohort we identified differences between the set of mutations that did not cause nullomer resurfacing and those that did. For instance, *TP53* was more frequently found to have nullomer resurfacing mutations relative to other cancer genes, and those nullomer resurfacing mutations were primarily missense mutations (**Figure 2a**). Interestingly, the variant allele frequency was higher in *TP53* for mutations that caused nullomer resurfacing (**Figure 2b**). Similar results were also obtained for other cancer genes such as *RYR2* (**Figure 2a**), indicating biases in the frequencies between mutations that did or did not cause nullomer resurfacing across patients.

We observed that for sixteen bp nullomers, more somatic mutations caused nullomer resurfacing than those that did not, across the top cancer genes (**Figure 2b-c**). For instance, 33% of patients had nullomer resurfacing mutations at *TP53*, whereas only 5% had mutations that did not cause nullomer resurfacing in the same gene (**Figure 2b-d**). We also found that the types of mutations that caused nullomer resurfacing in the most frequently mutated cancer genes were different from those that did not cause nullomer resurfacing and were primarily missense, nonsense and multi-hit mutations (**Figure 2b-d**). Thus, it can be inferred that in the vast majority of cases in which cancer genes are mutated, there is nullomer resurfacing associated with those mutations. Finally, significant differences were detected in the frequency of nullomer resurfacing between cancer types across kmer lengths (**Figure 2e**; **Supplementary Figure 6**). These nullomer signatures could be used in liquid biopsy as additional cancer biomarkers.

### Nullomer resurfacing at mutational hotspots

For the most mutated cancer genes, we compared the distribution and frequency of nullomer resurfacing mutations to those that did not cause nullomer resurfacing, across the length of each gene. Across the genetic pathways involved in cancer, we found that nullomer resurfacing mutations are more common than mutations that do not cause nullomer resurfacing (**Figure 3a**), which is consistent with the majority of mutations causing nullomer resurfacing at length sixteen (**Figure 1a**). When examining individual cancer genes, we observed that there were loci at which nullomers repeatedly resurfaced (**Figure 3b**; **Supplementary Table 1**) and these loci represented cancer driver events. In particular, oncogenes, such as *BRAF*, *PIK3CA* and *IDH1* showed individual nullomer resurfacing hotspots, whereas tumor suppressors such as *TP53* showed dispersed patterns of nullomer resurfacing across the gene body (**Figure 3b**). Thus, the characterization of nullomer resurfacing across individual cancer genes can enable their classification based on their origin and inform on the biological effect of a mutation.

**Figure 3:**
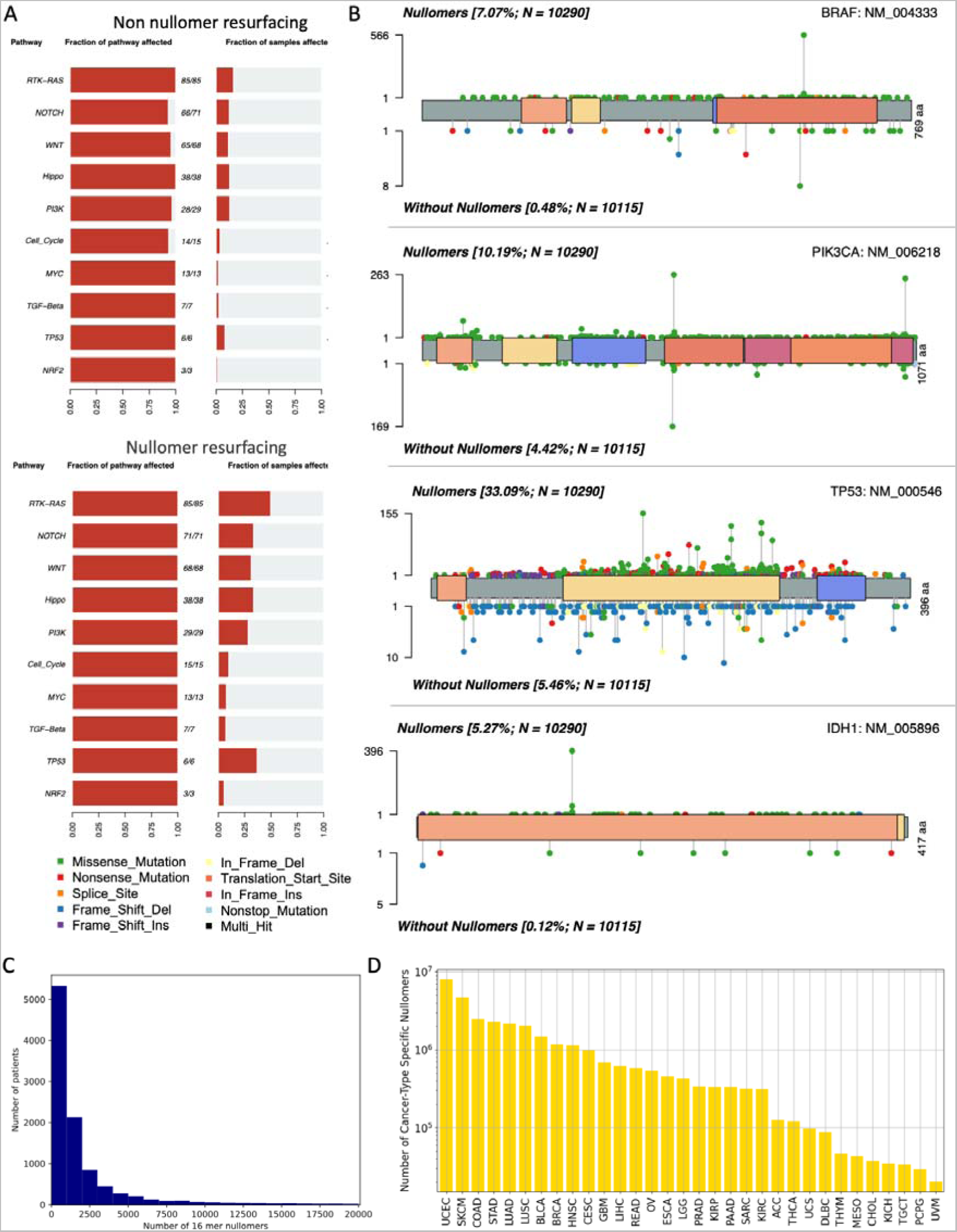
Identification of highly recurrent nullomers across cancer types and patients. **A.** Frequency with which genetic pathways were affected for mutations that do not cause or cause nullomer resurfacing. **B.** Lollipop plot displaying mutation distribution for nullomer resurfacing and non-nullomer resurfacing mutations. **C.** Number of patients in which each of the top sixteen-mer nullomers was detected. **D.** Number of cancer-type specific nullomers across all cancer types examined at length sixteen.

Nullomers that resurface recurrently across multiple cancer patients are more likely to be predictive of the tissue of origin of a cancer. We therefore examined how frequently each nullomer resurfaced in multiple patients across all the considered cancer types or at individual cancer types. We report that most nullomers are not recurrent; however, a small subset can be detected with high frequency across cancer patients (**Figure 3c**). We also showed that the most recurrently resurfacing nullomers are primarily found at a single cancer gene within a particular locus and primarily involve known driver mutations (**Table 1**). For instance, the most recurrently observed nullomer was found at *BRAF* across 5.5% of cancer patients, while other top resurfacing nullomers were found at individual loci in *IDH1*, *PIK3CA*, *KRAS* and *TP53* (**Table 1**), all of which are known cancer genes.

**Table 1:**
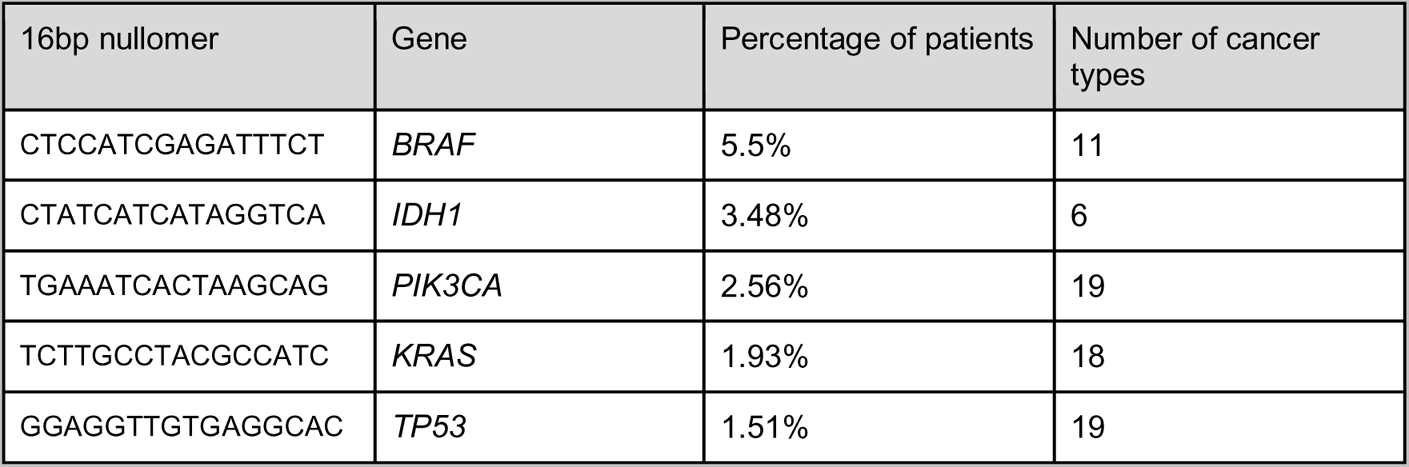
Selection of cancer genes and the corresponding most recurrently resurfacing 16bp nullomers across cancer types and patients. Selection is based on locus-specific nullomers.

Interestingly, we identified a second set of highly recurrent nullomers, which are observed at multiple cancer genes (**Table 2**). The top recurrent nullomers observed are found in clusters of paralogous genes. For instance, “CTCCAGTGTGAGTTAT” was found to resurface across 34 genes, of which most were zinc-finger genes, whereas “GTTGTTCTCGCGGACA” was found in 13 genes, all of which were different members of the Protocadherin Beta gene family. Therefore, highly recurrent nullomers can be identified across WES tumor samples and can be potentially utilized for the early detection of cancer with liquid biopsies.

**Table 2:**
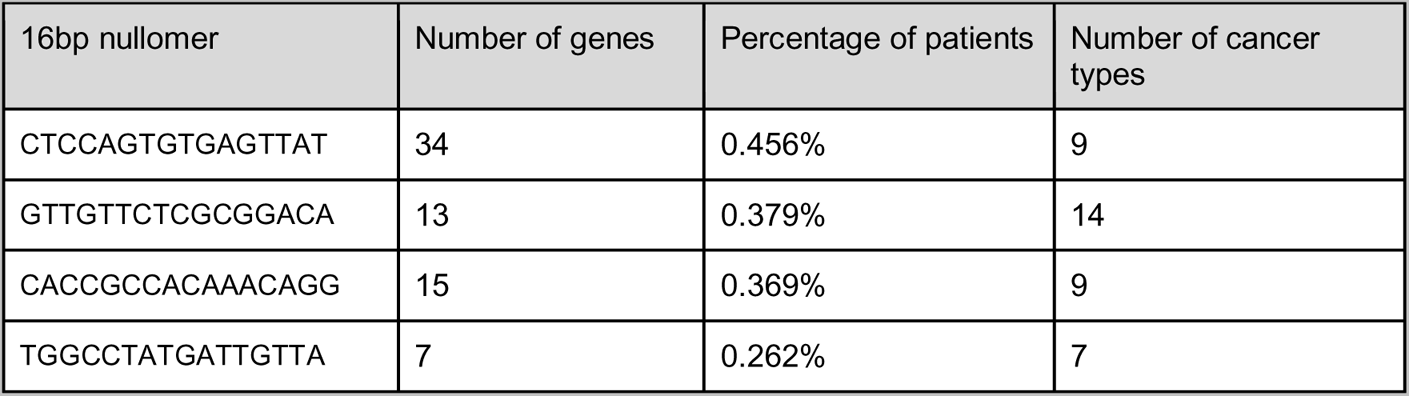
Selection of cancer genes and the corresponding most recurrently resurfacing 16bp nullomers across cancer types and patients. Selection is based on nullomers detected across multiple loci.

### Identification of cancer-type specific nullomers

We were also interested to investigate if certain nullomers appear in individual cancer types but are otherwise absent from all other cancer types and are, therefore, cancer-type specific. We identified cancer-type specific nullomers across all of the cancer types examined (**Figure 3d**), with the highest number of cancer-type specific nullomers being observed in uterine corpus endometrial carcinoma (UCEC), SKCM and colorectal adenocarcinoma (COAD), three of the cancer types with the highest mutational burden. We found that at longer kmer lengths the number of cancer-type specific nullomers being identified increased (**Supplementary Figure 7**). We also examined the recurrency of each nullomer at each tissue to investigate how frequently recurrent nullomers were shared between tissue types. We observed that nullomers that are highly recurrent in a tissue, are more likely to be shared between multiple tissue types (**Figure 4a**) and these likely reflect cancer driver events that occur across tissue types.

**Figure 4:**
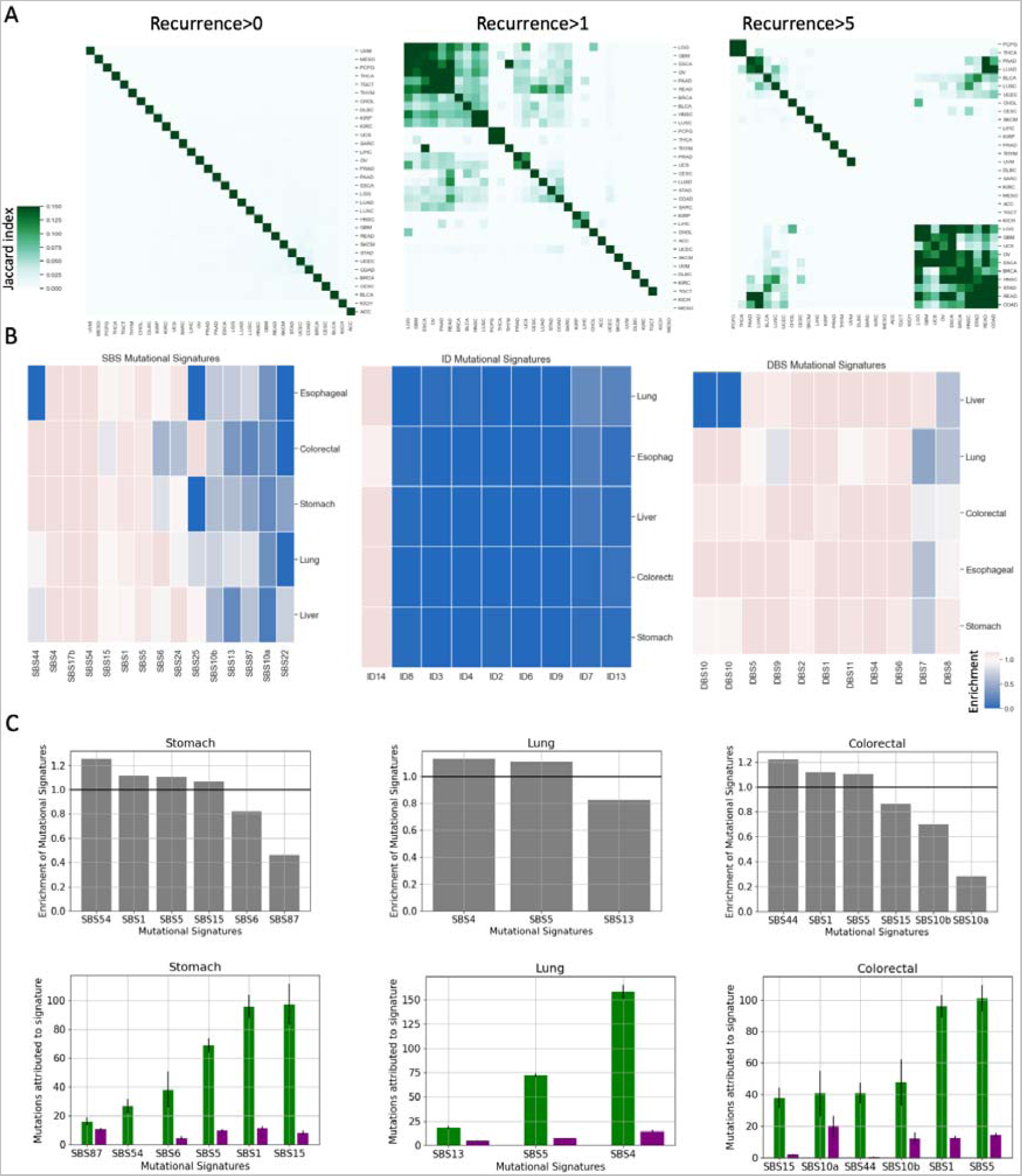
Mutational signatures that are linked to nullomer resurfacing across multiple cancer types. **A.** Hierarchical clustering presents which types of cancer have most common fifteen-mer nullomers. **B.** Enrichment of mutational signatures for mutations that cause nullomer resurfacing relative to mutations that do not cause nullomer resurfacing across five cancer types using sixteen bp nullomer length. **C.** Mutational signature enrichment patterns for SBS mutations in stomach, lung and colorectal cancers using sixteen bp nullomer length.

### Mutational signature analysis of nullomer-resurfacing mutations

Mutational signatures represent mutational processes that are operative during cancer development. We were interested to understand which mutational processes are most likely to cause nullomer resurfacing. We therefore examined if the mutational signatures that we detect in nullomer resurfacing mutations differ from those that do not cause nullomer resurfacing.

For this analysis we focused on five cancer types, namely lung, liver, esophageal, colorectal and stomach cancers. We fitted the mutational signatures for each sample separately for nullomer resurfacing and non-resurfacing mutations. For substitution mutations, we found that certain mutational signatures were more likely to cause nullomer resurfacing than others (**Figure 4b-c**; **Supplementary Figure 8**). For instance, we observed that SBS1 and SBS5 were more likely to cause nullomer resurfacing, which are signatures that correlate with age, and these findings were consistent across all the cancer types examined. In contrast, SBS87 which is associated with chemotherapy and SBS10a and SBS10b, which are associated with polymerase epsilon exonuclease domain mutations, were less likely to cause nullomer resurfacing.

We also performed the same analysis for doublet base substitutions (DBS) and indel mutational signatures. Our findings suggested that the indel signature ID14, which has an unknown etiology, is the only signature that is enriched at nullomer resurfacing mutations across the cancer types examined. In contrast, mutational signatures associated with defective mismatch repair are depleted of nullomer resurfacing mutations across cancer types (**Figure 4b**). For doublet substitutions, DBS2, DBS9 and DBS4 are enriched for nullomer resurfacing mutations. These findings show that the likelihood of nullomer resurfacing can be strongly influenced by the mutational signature across cancer types.

### Identification of clinically actionable nullomers

OncokB is a precision oncology database that incorporates clinically actionable targets, across different levels of clinical evidence (Chakravarty et al. 2017). We annotated somatic mutations using OncoKB and examined the frequency of nullomer resurfacing mutations at each level across the clinically determined levels, relative to the frequency of mutations that did not cause nullomer resurfacing. First, we compared nullomer resurfacing mutations and mutations that did not cause nullomer resurfacing for their functional consequence. We found that nullomer resurfacing mutations were more likely to cause gain of function, loss of function or switch of function, than mutations that did not cause nullomer resurfacing (**Figure 5a**). These results indicate that nullomer resurfacing cancer mutations are more likely to be functionally relevant. Next, we examined the frequency of nullomer resurfacing mutations across the OncoKB annotated therapeutic levels, to investigate if nullomers are enriched in therapeutically relevant mutations. We demonstrated that across the examined categories, mutations that caused nullomer resurfacing were significantly more likely to have therapeutic implications relative to mutations that did not cause nullomer resurfacing (**Figure 5b**). Considering the importance of the nullomer-resurfacing enrichment patterns of mutations for which there are Food and Drug Administration (FDA) approved drugs (Level 1), we detected all the clinically relevant nullomers that resurface from these mutations (certain of which are highlighted in **Supplementary Table 2**).

**Figure 5:**
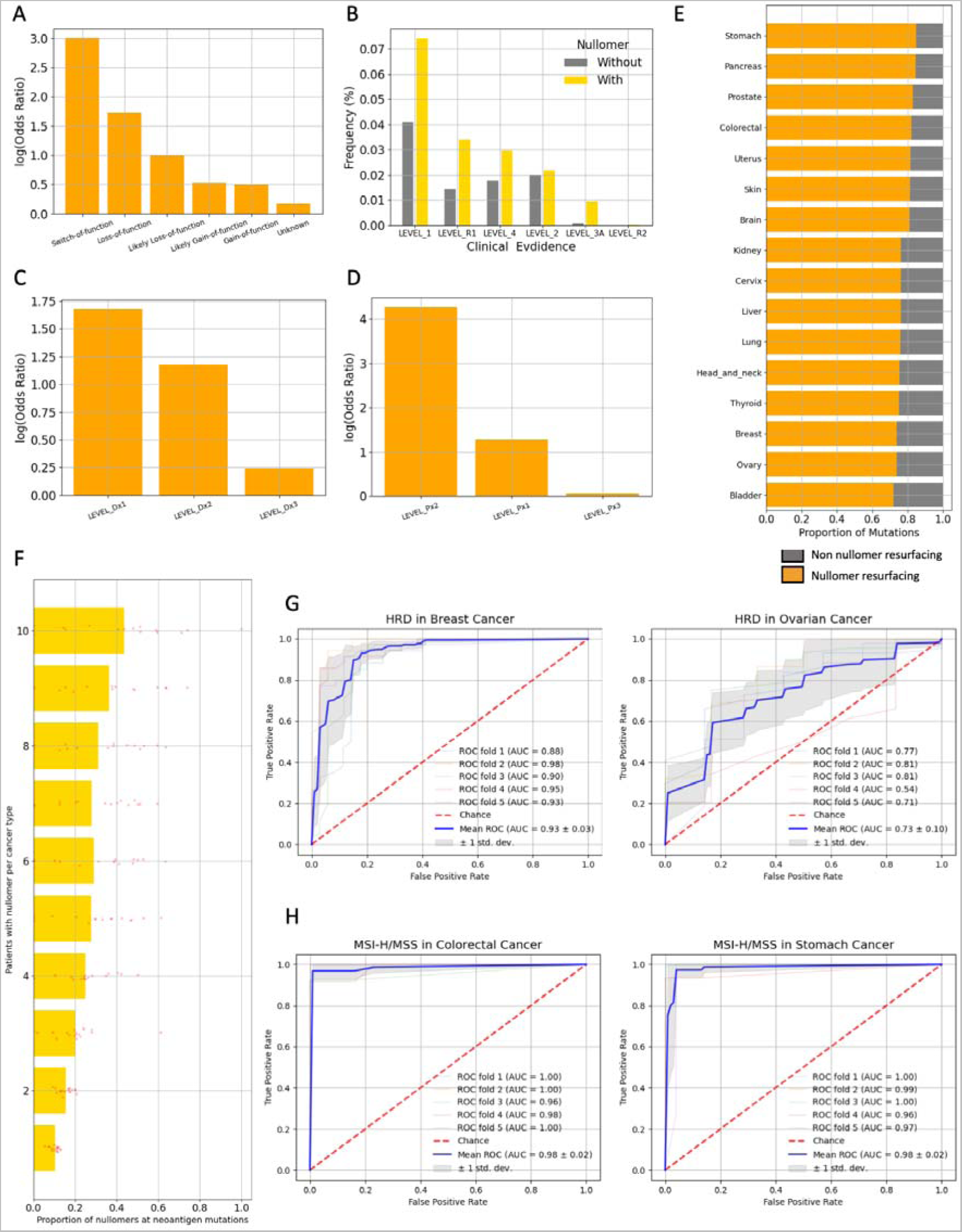
Nullomer resurfacing at loci that are clinically actionable targets. **A.** Functional consequence of nullomer resurfacing and non-nullomer resurfacing mutations. **B.** Therapeutic implication mutations at mutations that cause nullomer resurfacing and those that do not. **C.** Mutations with diagnostic importance and their enrichment at nullomer resurfacing mutations. **D.** Mutations with prognostic importance and their enrichment at nullomer resurfacing mutations. **E.** Nullomer resurfacing at known neoantigen generating mutations across multiple cancer types. **F.** Association between the recurrence of nullomer resurfacing in individual cancer types and its likelihood of being derived from a neoantigen generating mutation. Every dot represents a cancer type. **G.** HRD classification using nullomers in breast and ovarian cancers. **H.** MSI-H/MSS classification using nullomers in colorectal and stomach cancers.

We also examined if mutations that caused nullomer resurfacing were more likely to be classified as prognostic or diagnostic. For diagnosis, levels Dx1, Dx2, and Dx3 represent evidence that is required for diagnosis, supports diagnosis and is in investigational diagnosis, respectively. For prognosis, levels Px1 and Px2 refer to FDA and/or professional guideline-recognized biomarkers based on well-powered and small studies, respectively, whereas level Px3 refers to indication that is based on clinical evidence derived from well-powered studies. We observed that nullomer resurfacing mutations were significantly more likely to be classified as diagnostic (**Figure 5c**) and as prognostic (**Figure 5d**). For Px1 FDA and/or professional guideline-recognized targets we presented the generated nullomers (certain of which are highlighted in **Supplementary Table 3–4**). Our results provide evidence for the utility of nullomer identification for cancer detection, prognosis and diagnosis, as well as for precision oncology purposes.

Recurrent fusion events are commonly observed across multiple cancer types (Mertens et al. 2015). We derived cancer gene fusion events from ChimerDB 4.0 (Jang et al. 2020) and identified the nullomers resurfacing from them. We report that gene fusion events are also a source of nullomer resurfacing across gene fusion events, including highly recurrent events such as EML4-ALK and BCR-ABL1 among others (**Supplementary Table 5**).

The detection of neoantigen targets is critical for the development of personalized cancer immunotherapies (Lang et al. 2022). Therefore, identification of neoantigen targets in liquid biopsy samples offers an attractive target. We derived neoantigens for fifteen cancer types from TSNAdb (Wu et al. 2022) and examined if we can identify nullomers resurfacing from mutations that cause neoantigen formation (certain of which are highlighted in **Supplementary Table 6**). Across the cancer types studied, the largest proportion of neoantigen generating mutations causing nullomer resurfacing was observed for stomach and pancreatic cancers, whereas the lowest proportion was observed in bladder cancer (**Figure 5e**; **Supplementary Figure 9**). We also examined the frequency with which the neoantigen associated nullomers appeared as a function of nullomer recurrency. We observed that across cancer types, nullomer recurrence was linked to a higher likelihood of being a neoantigen target (**Figure 5f**). These results provide evidence for the utility of nullomers in the detection of neoantigens.

### Identification of DNA repair deficiencies with nullomers

Next, we investigated if we can identify common DNA repair deficiencies using nullomers in WES tumor samples. The HRD score is a measure of genomic instability that is used to detect homologous recombination deficiency (Telli et al. 2016). Here, we examined if we can separate samples with high HRD score (>42), which is used to define homologous recombination deficiency, from samples that do not have homologous recombination deficiency (Telli et al. 2016). We developed a nullomer-based classifier in two cancer types, namely ovarian and breast cancer, and showed that we can detect homologous recombination deficiency with AUC scores of 0.73 and 0.93 respectively, using nullomers (**Figure 5g**). The results were similar when also using 14bp and 15bp kmer length nullomers (**Supplementary Figure 10a-b**) and we also observed that a subset of nullomers were more informative than others in the classification models (**Supplementary Figure 11**). The lower performance in the case of ovarian cancer was attributed to the lower sample size, and when we generated a single classifier able to predict homologous recombination deficiency in both ovarian and breast cancer, the prediction performance improved substantially (AUC = 0.91; **Supplementary Figure 10c-e**).

Furthermore, mismatch repair deficiency can be used as a biomarker to guide treatment with immune checkpoint inhibitors (Yi et al. 2018). Here, we examined if we can detect High Microsatellite Instability (MSI-H) samples from microsatellite stable (MSS) samples. We observed that in both colorectal and stomach cancer, we accurately detected mismatch repair deficiency with nullomers (AUCs of 0.98 in both cancer types; **Figure 5h**; **Supplementary Figure 12**). These findings indicate that nullomers detected in WES can be used to stratify patients and to detect DNA repair deficiencies.

### Identification of nullomers in cfRNA for cancer detection

We examined if the identified nullomers can be used to detect cancer in liquid biopsies using cfRNA data. We performed our analyses using two datasets that encompassed lung, colorectal, stomach, esophageal and liver cancers as well as healthy controls (Zhu et al. 2021; S. Chen et al. 2022). For each sample, we identified the nullomers present for nullomer lengths between 14bp and 16bp and generated classification models to estimate our ability to detect cancer. The nullomers that we incorporated in this analysis were the top 100,000 most frequently resurfacing nullomers across all cancer types (**Figure 1**), serving as a general list of nullomers to detect multiple cancer types.

For the first dataset, which encompassed hepatocellular carcinoma (HCC) and healthy control data, we examined the frequency of nullomer resurfacing in cancer relative to controls (Zhu et al. 2021). We observed that the total counts of nullomers detected in cfRNA derived from liquid biopsies of HCC patients was significantly higher than for the healthy controls (**Welch Two Sample t-test, p-value<0.0001 across kmer lengths; Figure 6a**). Next, we examined if the size of the set of unique nullomers differed between the two groups and found consistent patterns (**Welch Two Sample t-test, p-value<0.0001 across kmer lengths; Figure 6b**). We also trained a machine learning model to examine if we can accurately detect HCC based on the nullomers identified in each sample. We generated a lasso logistic regression classification model which was able to detect cancer samples in all cases (AUC=1; **Figure 6c**). Results were highly consistent with different kmer lengths and we were able to accurately detect HCC also using fourteen (AUC=0.999) and fifteen (AUC=0.998) bp nullomer lengths (**Supplementary Figure 13a-b**). In addition to good discrimination between HCC and healthy samples, each model showed accurate probabilistic predictions as evidenced by a Brier score less than or equal to 0.02 (**Supplementary Figure 14a-c**). We also examined the top most informative features and observed that the most informative nullomers were found at liver cancer associated genes, including *FTH1*, *EEF2*, *TMSB10*, *ACTB* and the long non-coding RNA *MALAT* among others (**Table 3**). Our findings provide evidence for the utility of nullomer identification in cfRNA for cancer detection.

**Figure 6:**
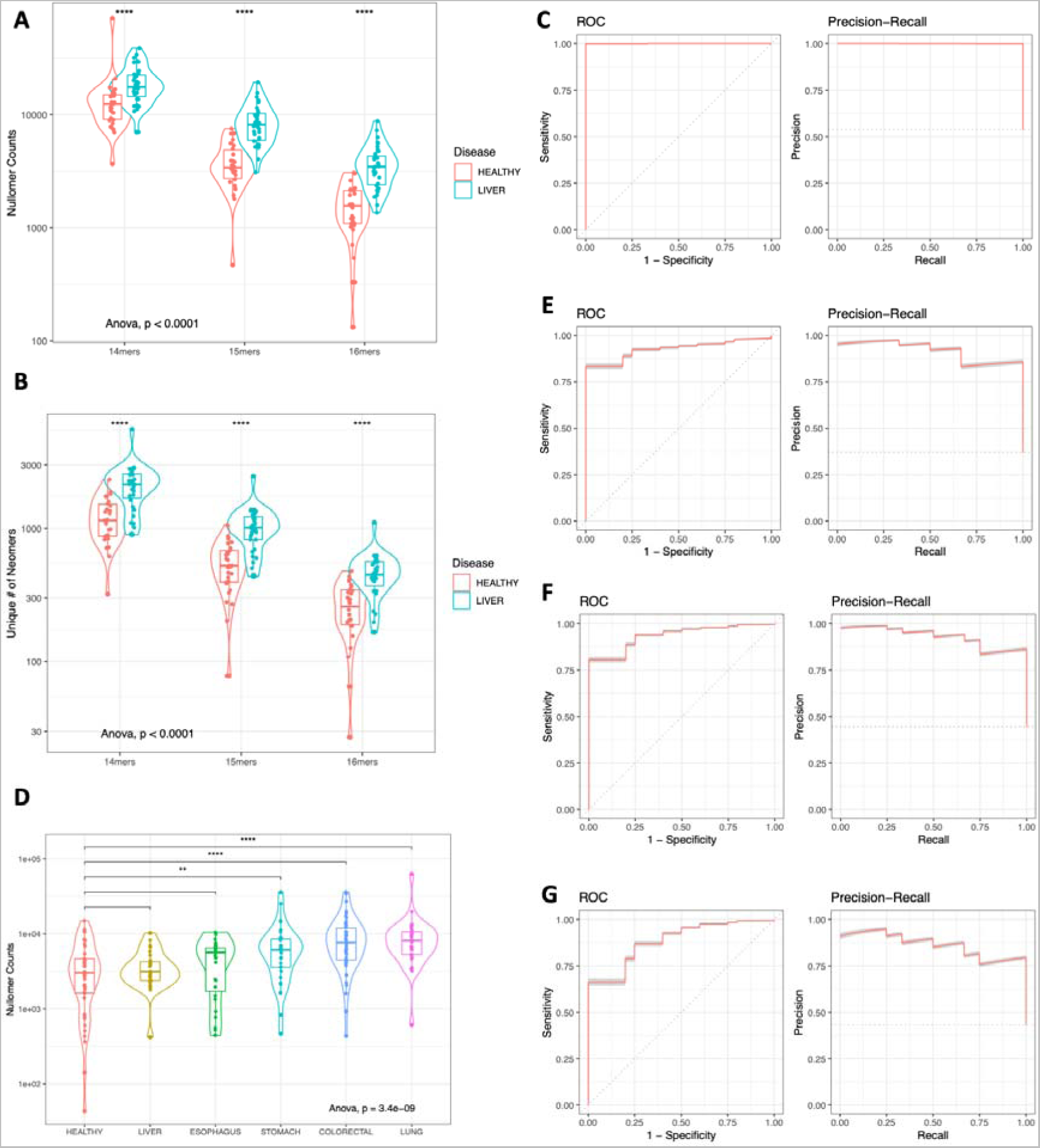
Identification of nullomers in cfRNA derived from liquid biopsy samples for early cancer detection. **A.** Counts of nullomers identified in healthy and HCC samples using 14mer, 15mer and 16mer nullomers. Samples are grouped by disease state. **B.** Number of unique nullomers identified in healthy samples and HCC. Results shown for 14mer, 15mers and 16mer nullomers. **C.** ROC curve and precision recall for liver cancer. **D.** Counts of nullomers identified in healthy and cancer samples for liver, esophageal, stomach, colorectal and lung cancers using 16mer nullomers. Cancer samples are grouped by cancer type. **E.-G.** ROC curve and precision recall for **E.** liver cancer, **F.** stomach cancer and **G.** lung cancer.

**Table 3:**
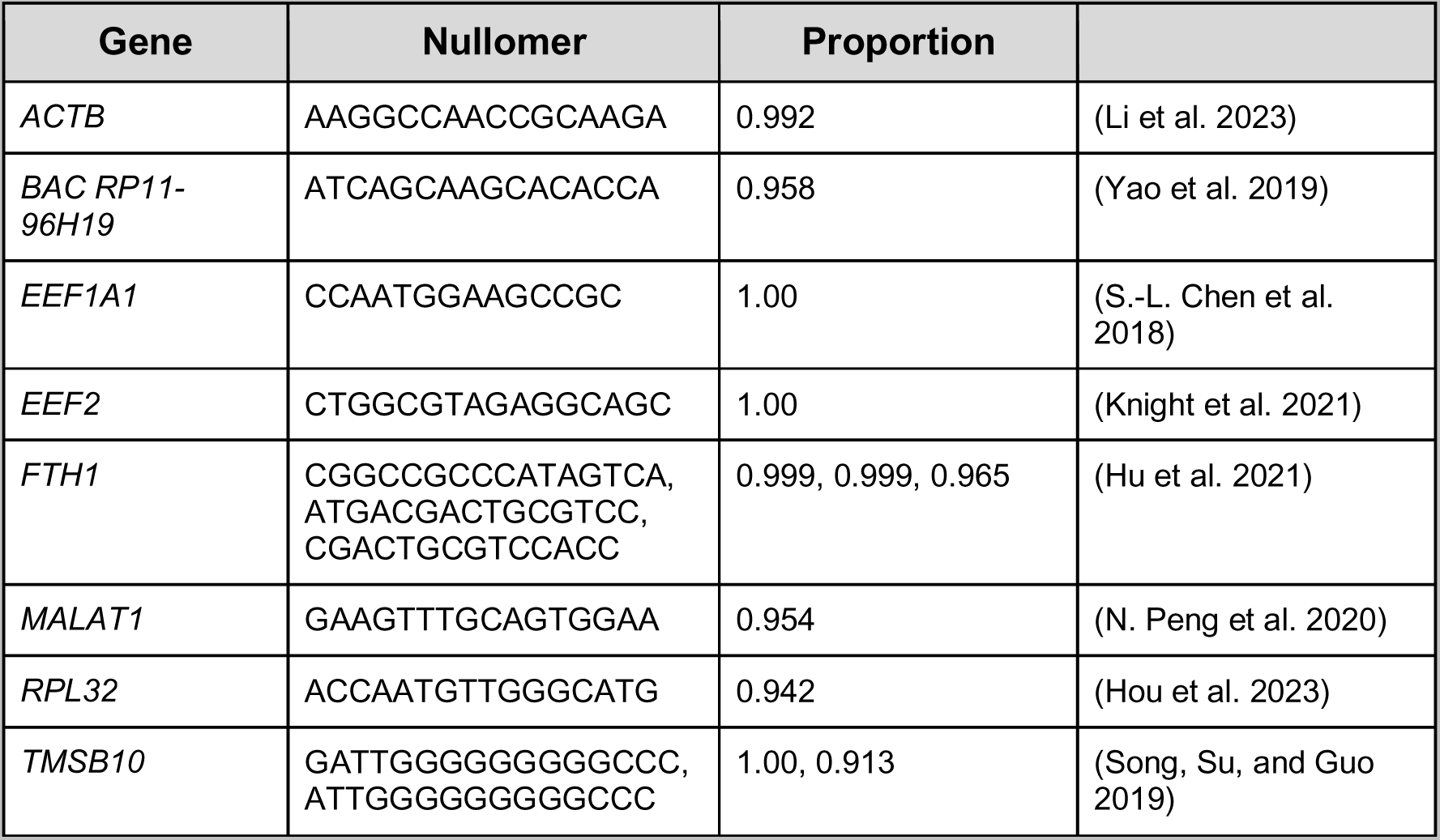
Selection of the most informative nullomers for detection of liver cancer.

Next, we examined a dataset that included liquid biopsy derived cfRNA data from liver, esophageal, stomach, colorectal and lung cancers as well as healthy control cfRNA data (S. Chen et al. 2022). We found that, on average, samples from each cancer type displayed more nullomer counts than the controls (**Figure 6d**, **Welch Two Sample t-test**, p-value=0.0001491), suggesting that nullomers can indeed be used to differentiate between cancer patients and healthy controls across disparate cancer types. Next, we also created lasso logistic regression classification models for cancer detection and examined their performance for each cancer type. The classification models for liver, stomach, and lung cancer had an AUC of 0.922, 0.927, and 0.877, respectively. The reported results indicated the models’ ability to accurately classify cancer and healthy samples across different cancer types (**Figure 6d-g**) with particularly high performance for stomach, thus revealing the potential of RNA nullomers to facilitate early cancer detection.

**Table 4:**
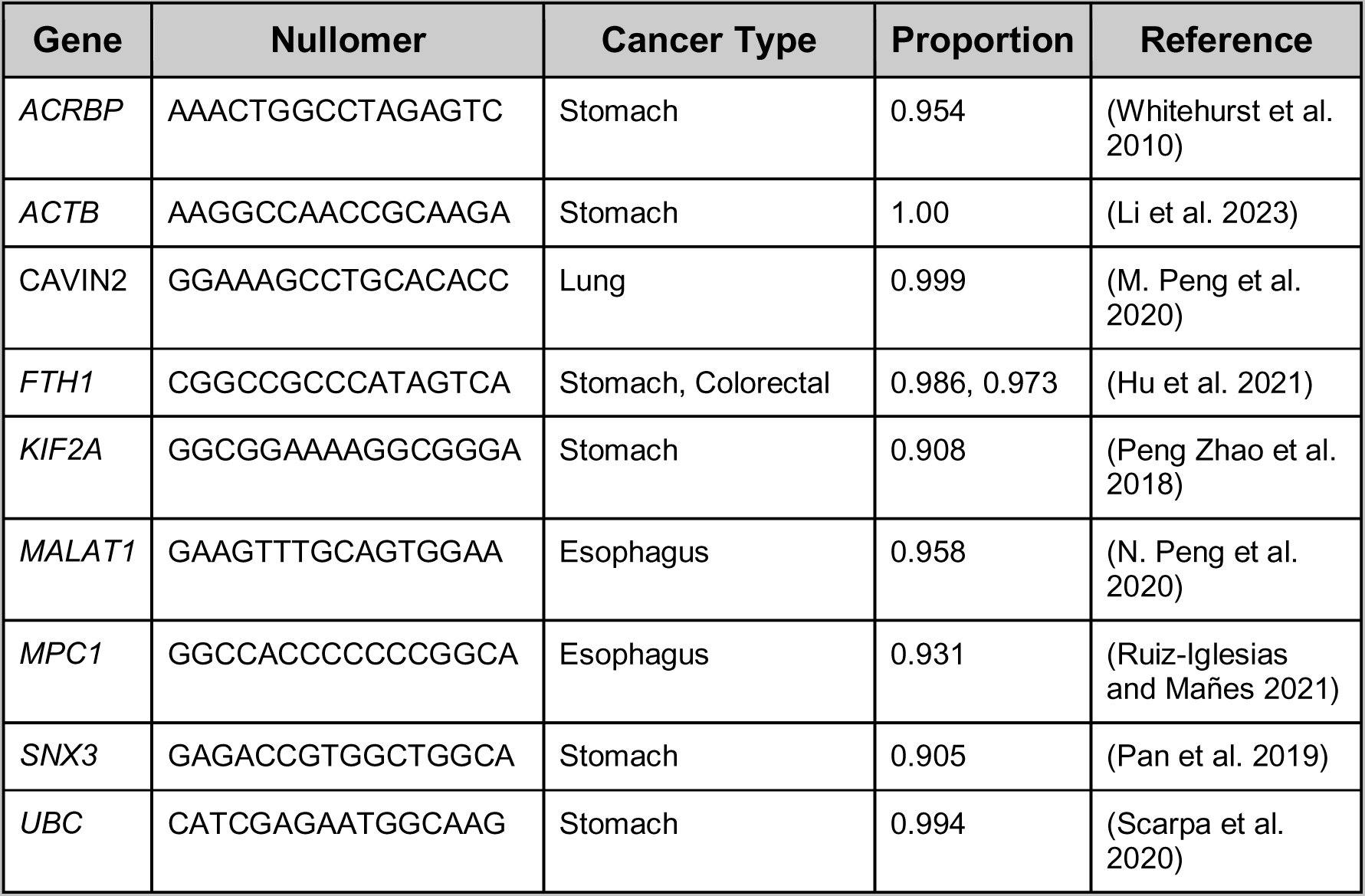
Selection of the most informative nullomers for detection of multiple cancer types including stomach, lung, colorectal, esophageal and liver cancers.

## Discussion

In this study, we have characterized nullomer-resurfacing across more than 10,000 WES tumors in 32 cancer types and investigated their utility as cancer biomarkers in liquid biopsies with cfRNA. The usage of a cfRNA-based cancer detection assay offers several advantages. For instance, the process by which tumor-derived RNA is introduced into the bloodstream likely exhibits differences from cfDNA, including its transfer with exosomes (Larson et al. 2021; Enderle et al. 2015). In addition, the usage of cfRNA in diagnostics can incorporate overall expression levels and dynamic expression changes, which can prove particularly important for tumors with lower mutational burden.

In contrast to the usage of nullomers in WGS tumor samples, in which the vast majority of the identified nullomers are non-coding and are passenger mutations, in WES we observe a substantial fraction of them being cancer drivers and actionable targets. Furthermore, we examine the mutational processes that are more likely to cause nullomer resurfacing and find that certain mutational signatures including aging mutational signatures are more likely to cause nullomer resurfacing. These findings have direct implications in improving clinical practice for cancer patients using a minimally invasive procedure, which remains an area of ongoing research (Ignatiadis, Sledge, and Jeffrey 2021).

We also provide evidence for the usage of nullomers in cfRNA for early cancer detection across multiple cancer types. In future work we plan to incorporate additional disparate cancer types to characterize the performance of our nullomer based approach between them. It will be of interest to directly compare the performance of predictive models using cfDNA and cfNA for the same patients as well as their integration into multi omics predictive models. Furthermore, as immunotherapies and personalized treatments are advancing, nullomer based cfRNA-based diagnosis could be coupled with the identification of neoantigens for personal cancer vaccine development or other patient-tailored therapies.

Therefore, in future work we envision an integrated setting in which we can use nullomers across the stages of cancer care including cancer detection, diagnosis and treatment choice. Finally, cfRNA biomarkers can be combined with DNA-based, protein-based and other cancer biomarkers to improve and advance the early diagnosis of cancer.

## Methods

### Mutation dataset

Whole exome sequencing mutation data from tumor samples with matched controls were downloaded from https://api.gdc.cancer.gov/data/1c8cfe5f-e52d-41ba-94da-f15ea1337efc for over 10,000 whole exome sequencing tumor samples spanning 32 cancer types, from The Cancer Genome Atlas. Throughout the study, the GRCh37 reference human genome was used unless otherwise stated.

### Nullomer resurfacing from somatic mutations

Nullomers were identified as previously described in (Georgakopoulos-Soares, Yizhar-Barnea, et al. 2021). Nullomer resurfacing was performed for kmer lengths of 12-16bp for each somatic mutation across cancer patients and tumor types. Somatic mutations were separated into nullomer resurfacing and mutations that did not cause nullomer resurfacing. Maftools was used for the analysis of somatic mutations across cancer genes and at individual loci of specific cancer genes.

### Mutational signature analysis

Mutational signature analysis was performed using the SigProfilerExtractor package (Islam et al. 2022). Mutational signatures were fitted to the mutations of each tumor sample separately for mutations that caused nullomer resurfacing and mutations that did not cause nullomer resurfacing. The mutational signature enrichment was defined as the ratio of the proportion of mutations attributed to the signature over all the mutations for nullomer resurfacing mutations over the proportion of mutations attributed to the signature over all the mutations for mutations that did not cause nullomer resurfacing.

### Identification of cancer-type specific nullomers

Tumor types were clustered based on the proportion of nullomers shared. Cancer-type specific nullomers represented nullomers that resurfaced in at least one patient within a cancer type and which were absent from every patient across all other cancer types.

### Characterization of nullomers as actionable targets

Clinically actionable mutations were derived from OncoKB using the API to annotate somatic mutations with oncokb-annotator (Chakravarty et al. 2017). For each mutation we examined if it caused nullomer resurfacing and stratified by functional consequence and by FDA, prognostic, diagnostic and therapeutic levels, for every kmer length up to and including 16bp.

Neoantigens for fifteen cancer types were obtained from TSNAdb (Wu et al. 2022) and each neoantigen generating mutation was examined for nullomer resurfacing. Nullomer lengths up to 16bp long were examined.

### Identification of nullomers in fusion events

Fusion events were obtained from ChimerDB 4.0 (Jang et al. 2020). For each fusion event, the nullomers that resurface were derived for nullomer lengths up to 16bp.

### Identification of DNA repair enzyme vulnerabilities

For the detection of homologous recombination deficiency we utilized the MRD score as calculated from (Thorsson et al. 2018). Samples with HRD score > 42 were compared against the same number of samples with the lowest HRD scores and a classifier was developed to differentiate the two groups, independently for each cancer type. The feature space was derived from the somatic mutations at sixteen bps nullomers. The same analysis was repeated for fifteen and fourteen bps nullomers. The training set constituted 80% of the samples and the classification process used a random forest classification method with five-fold cross validation.

For the detection of mismatch repair deficiency through microsatellite instability, we used the classification of samples in MSI-H and MSS from (Liu et al. 2018). Nullomers up to and including 16bp long, were used for the development of classification models. The training set constituted 80% of the samples and the classification process used a random forest classification method with five-fold cross validation. The training and performance of the models was independently tested in colorectal and stomach cancers.

### cfRNA dataset processing

Liquid biopsy cfRNA fastq files were downloaded (Zhu et al. 2021; S. Chen et al. 2022). Sequences containing the top 100,000 resurfacing nullomers (12bp to 16bp) across cancer types were extracted from each sample’s respective fastq files with BBTools bbduk.sh (Bushnell, Rood, and Singer 2017). The resulting reads were then trimmed with Trim Galore (Krueger et al. 2023) to remove poor quality bases and adapter sequences. The reads were then filtered with BBTools seal.sh (Bushnell 2014) to remove common microbial contaminants, UniVec, ERCC spike-in, and ribosomal sequences. The remaining reads were subsequently deduplicated with BBTools dedupe.sh (Bushnell 2014). Aligning was done with BBTools bbmap.sh (Bushnell 2014) with stringent parameters (minid=0.9 kfilter=25) against a custom genome including GRCh38, SILVA SSU Ref NR 99, and common human viruses. Jellyfish (Marçais and Kingsford 2011) was used to count occurrences of each nullomer in the aligned SAM files.

### Classification model to detect cancer patients from cfRNA

A count matrix of samples by nullomers was used as the starting input for each model. Any counts less than or equal to two were set to zero to decrease false positive counts. Nullomers which had a sum of counts across samples less than 10 were removed. The count matrix was then CPM normalized with edgeR (Robinson, McCarthy, and Smyth 2010). Samples for each matrix contained healthy samples and samples of a specific cancer type. The R caret package (Kuhn 2008) was used to tune the lambda parameter of an L1 regularized logistic regression model across twenty values between zero and one. 10 fold cross-validation was repeated 100 times to detect the best model and evaluate its stability. Models were calibrated with the val.prob function in the rms R package. Models which showed a sigmoid curve between predicted probability and actual probability were then recalibrated with Platt scaling (Platt 1999). The R precrec package (Saito and Rehmsmeier 2017) was used to generate the Precision-Recall and ROC curves based on the repeated cross-validation predictions. Feature importance was assessed using the R glmnet package (Friedman, Hastie, and Tibshirani 2010) to perform 10 fold cross-validation repeated 100 times with the previously ascertained lambda parameter. Nullomers with non-zero coefficients were tracked across each of the 1000 models. Nullomers which occurred in over 90% of the models were deemed important and stable across models.

## Data Availability

The code for this work can be found at:
https://github.com/Georgakopoulos-Soares-lab/cfRNA-nullomer-analysis

## Supplementary Material

**Supplementary Figure 1:**
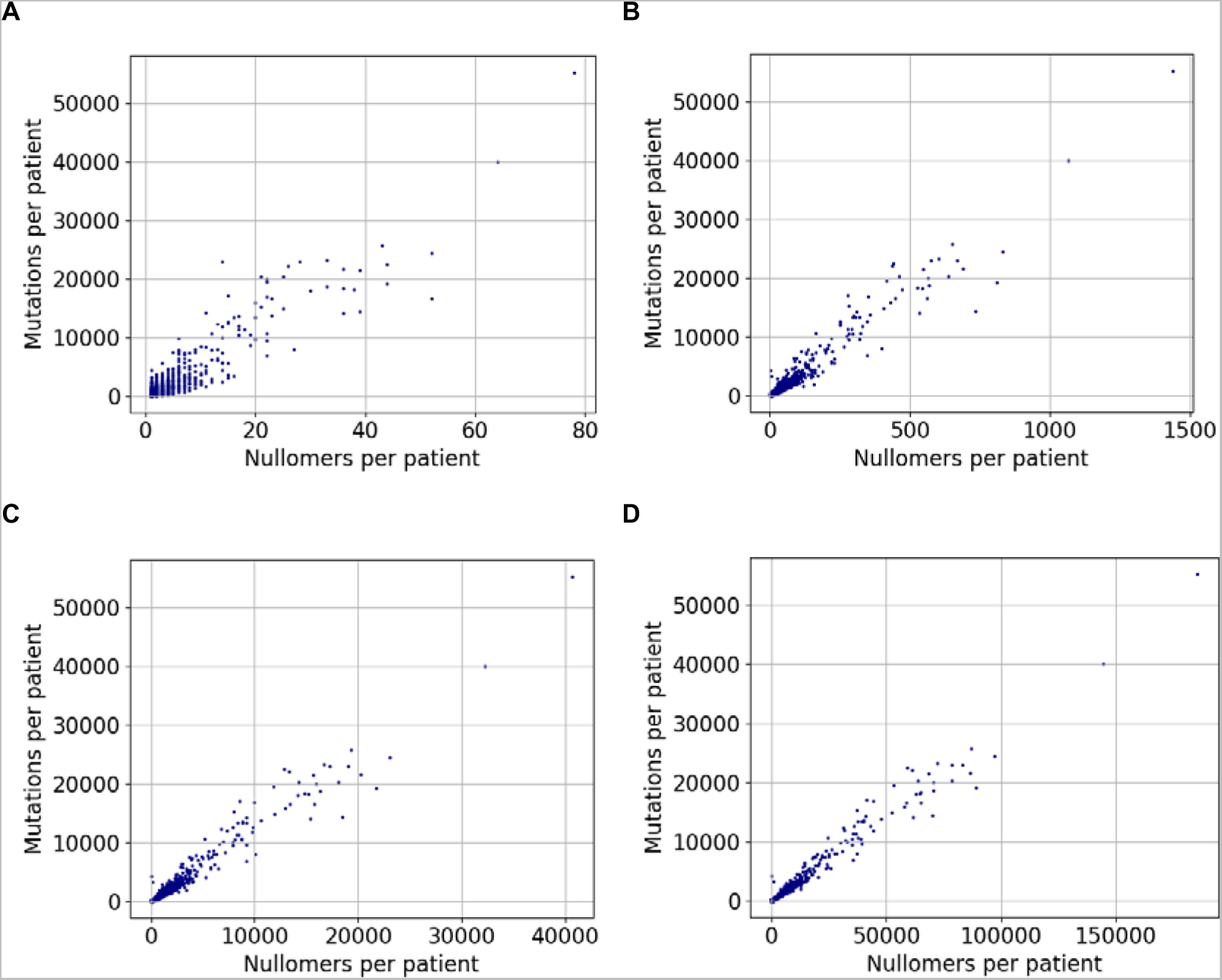
Association between the number of protein-altering mutations and the number of nullomers that resurface per patient. Results shown for **A.** twelve-mer nullomers and **B.** thirteen-mer nullomers, **C.** fourteen-mer nullomers, **D.** fifteen-mer nullomers. In A-D every dot represents a patient sample.

**Supplementary Figure 2:**
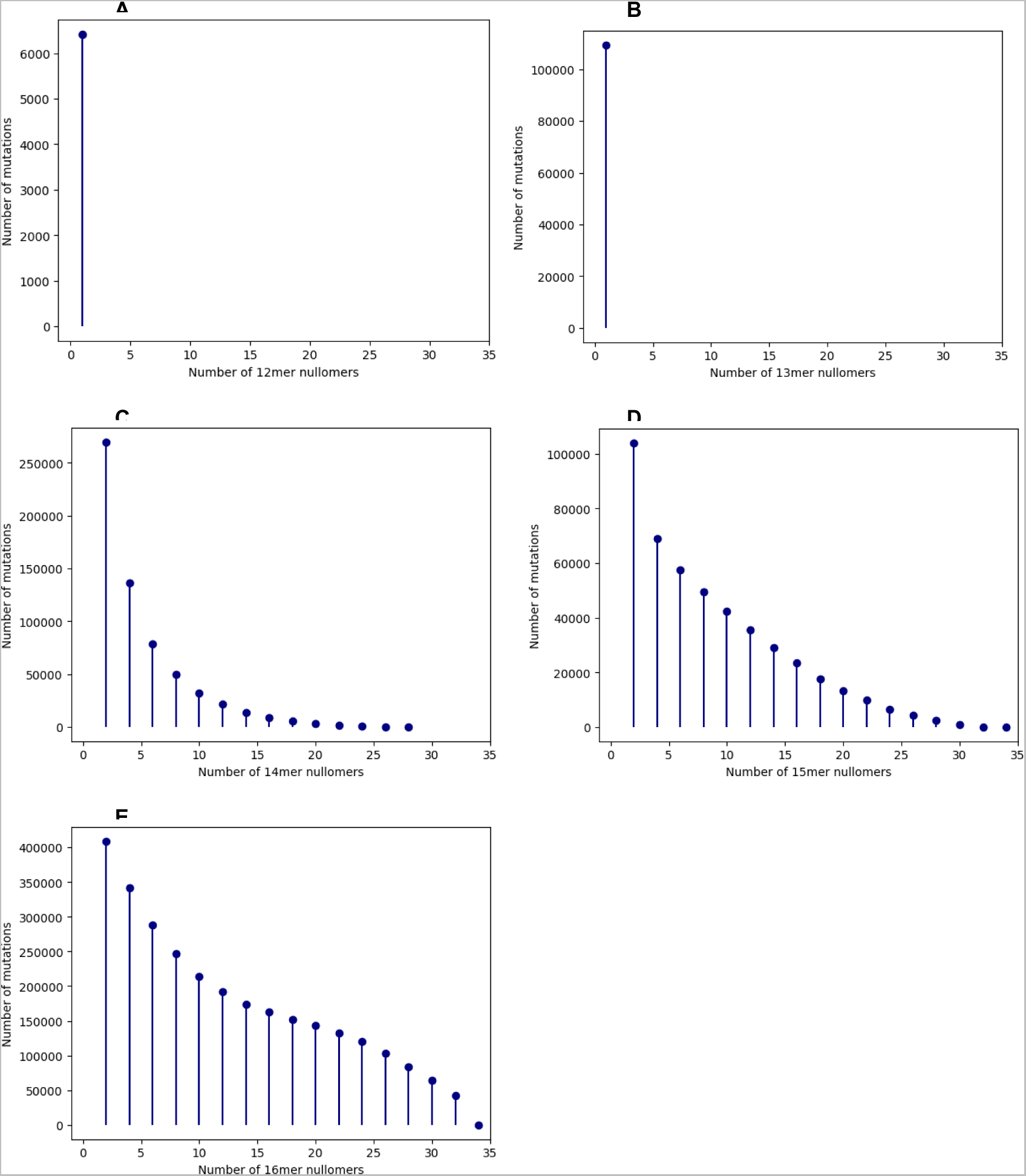
Number of nullomers produced from every mutation. **A.** twelve-mer nullomers and **B.** thirteen-mer nullomers, **C.** fourteen-mer nullomers, **D.** fifteen-mer nullomers. **E.** sixteen-mer nullomers.

**Supplementary Figure 3:**
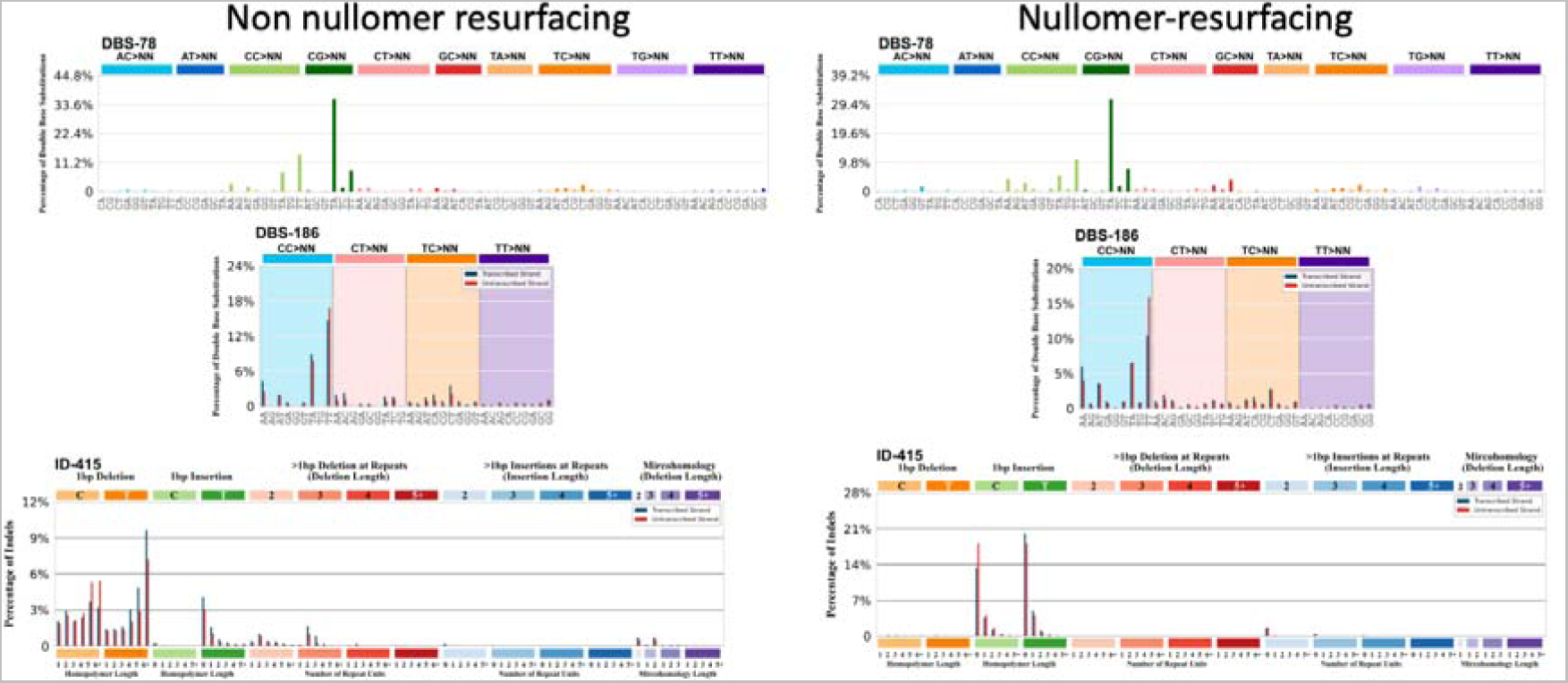
Mutational profile of doublet base substitutions (DBS) and indel (ID) mutations. Results shown for mutations that do not cause nullomer resurfacing (left) or those that cause nullomer resurfacing (right).

**Supplementary Figure 4:**
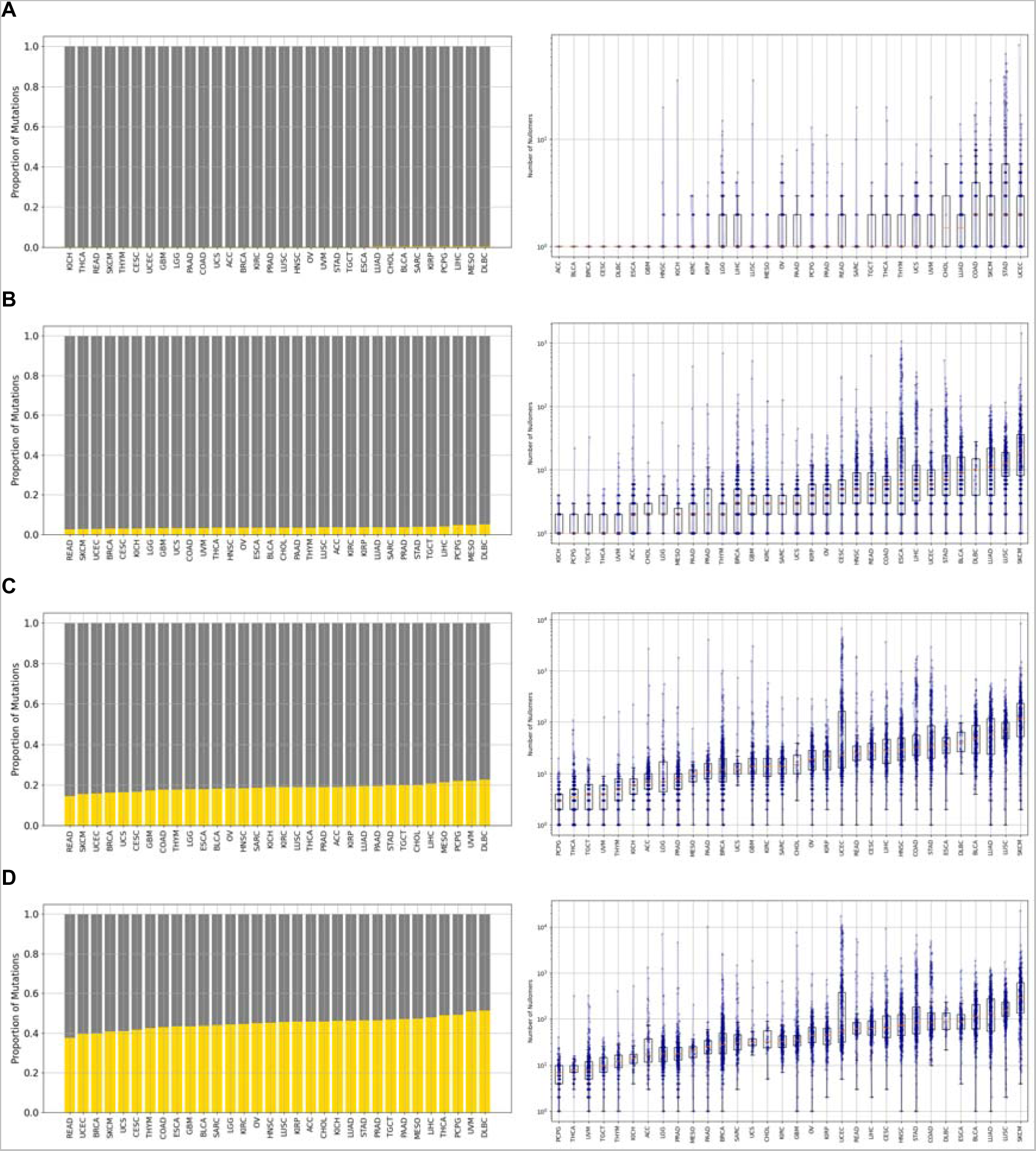
Proportion of mutations causing nullomer resurfacing across cancer types. **Results shown for: a.** 12mers, **B.** 13mers, **C.** 14mers and **D.** 15mers.

**Supplementary Figure 5:**
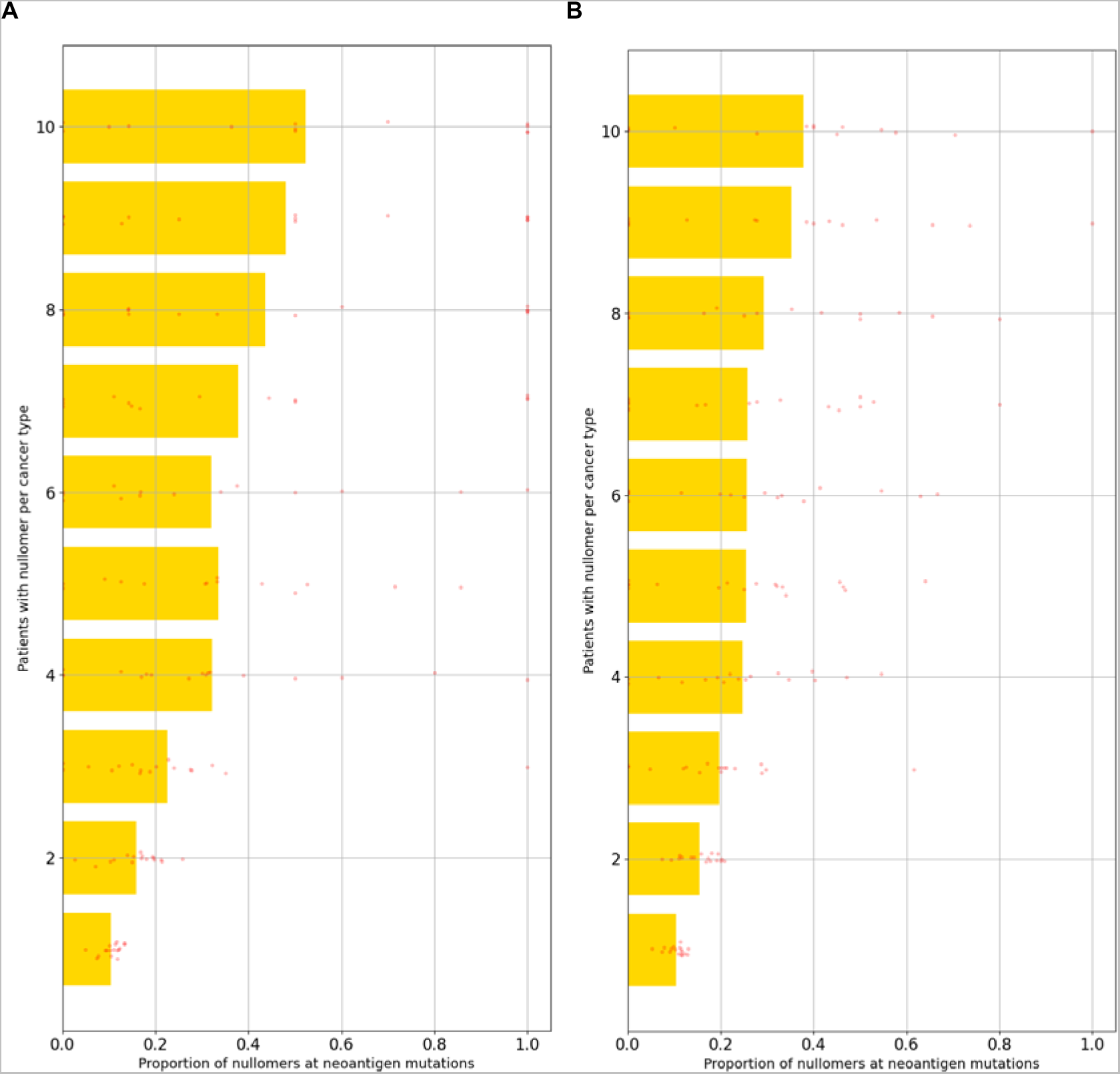
Association between the recurrence of nullomers in individual cancer types and the likelihood of them being derived from a neoantigen generating mutation. Results shown for **A.** 14bp and **B.** 15bp nullomer lengths.

**Supplementary Figure 6:**
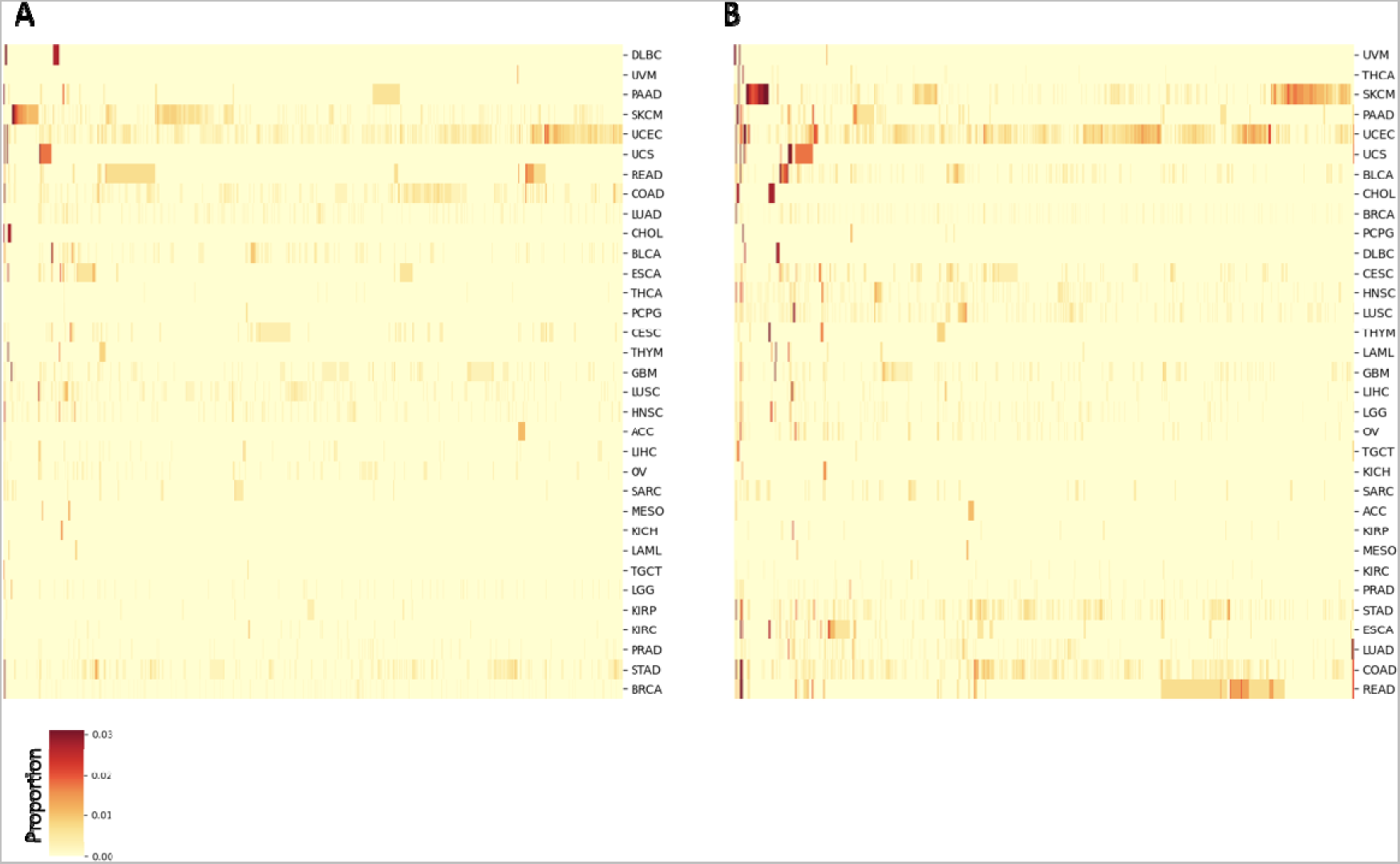
Proportion of patients in which each of the top 10,000 nullomers is found in for: A. 14bp nullomers, B. 15bp nullomers.

**Supplementary Figure 7:**
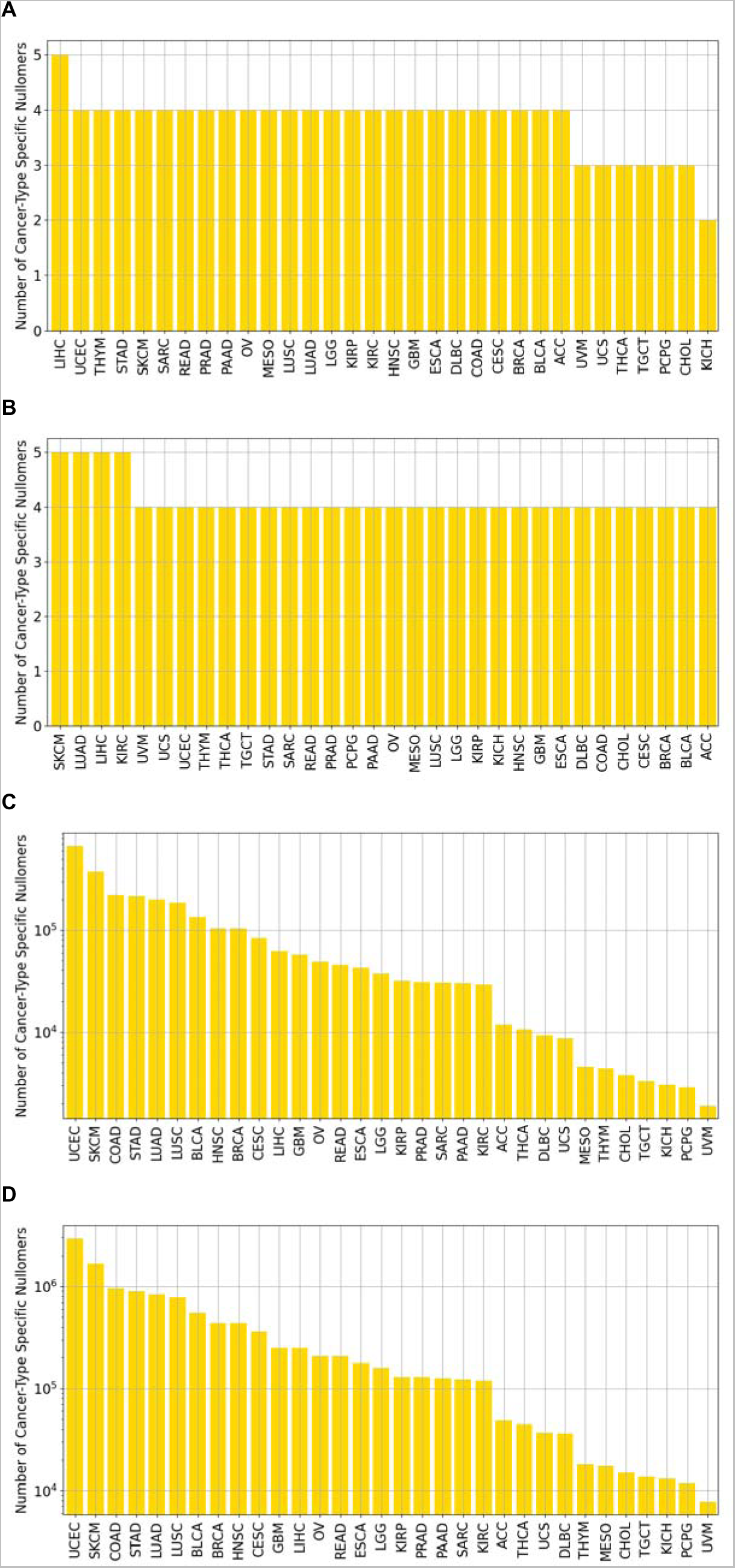
Identification of cancer-type specific nullomers. **A.** Percentage of unique **A.** 12-mer, **B.** 13-mer, **C.** 14-mer and **D.** 15-mer nullomers per cancer type.

**Supplementary Figure 8:**
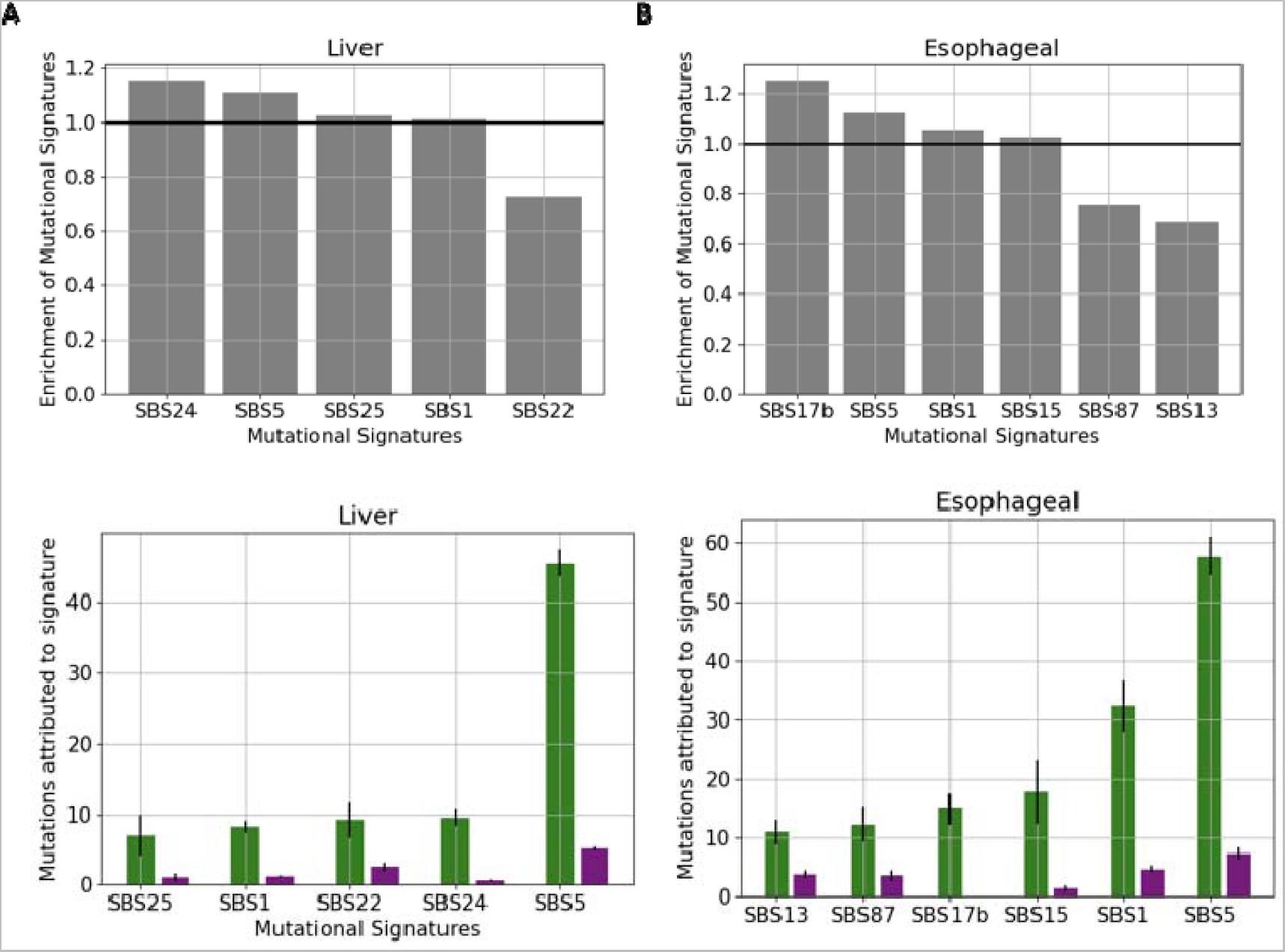
Mutational signature profile for mutations that either cause or do not cause nullomer resurfacing across A. Liver, B Esophageal cancer.

**Supplementary Figure 9:**
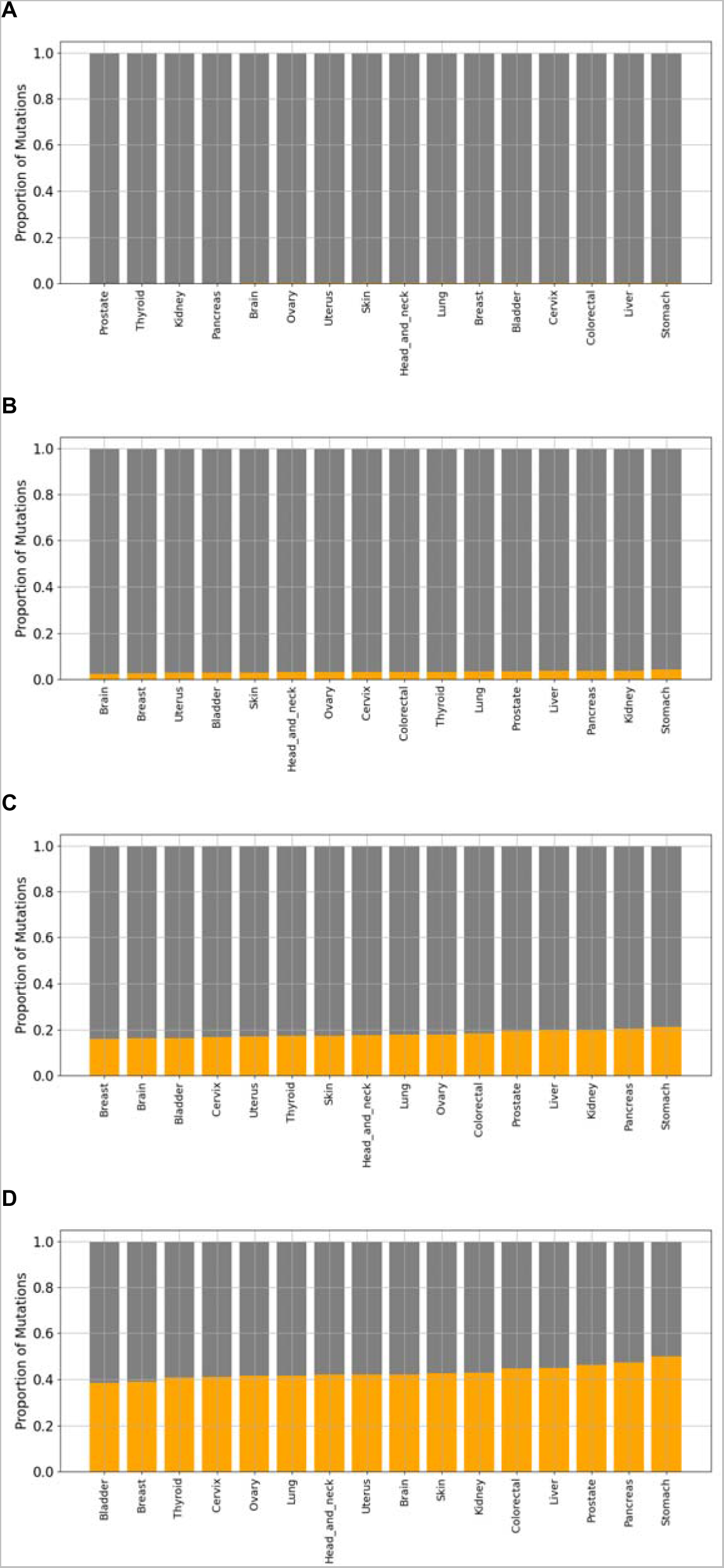
Neoantigen generating mutations cause the resurfacing of nullomers. Results shown for: **A.** 12-mers, **B.** 13-mers, **C.** 14-mers, **D.** 15-mers.

**Supplementary Figure 10:**
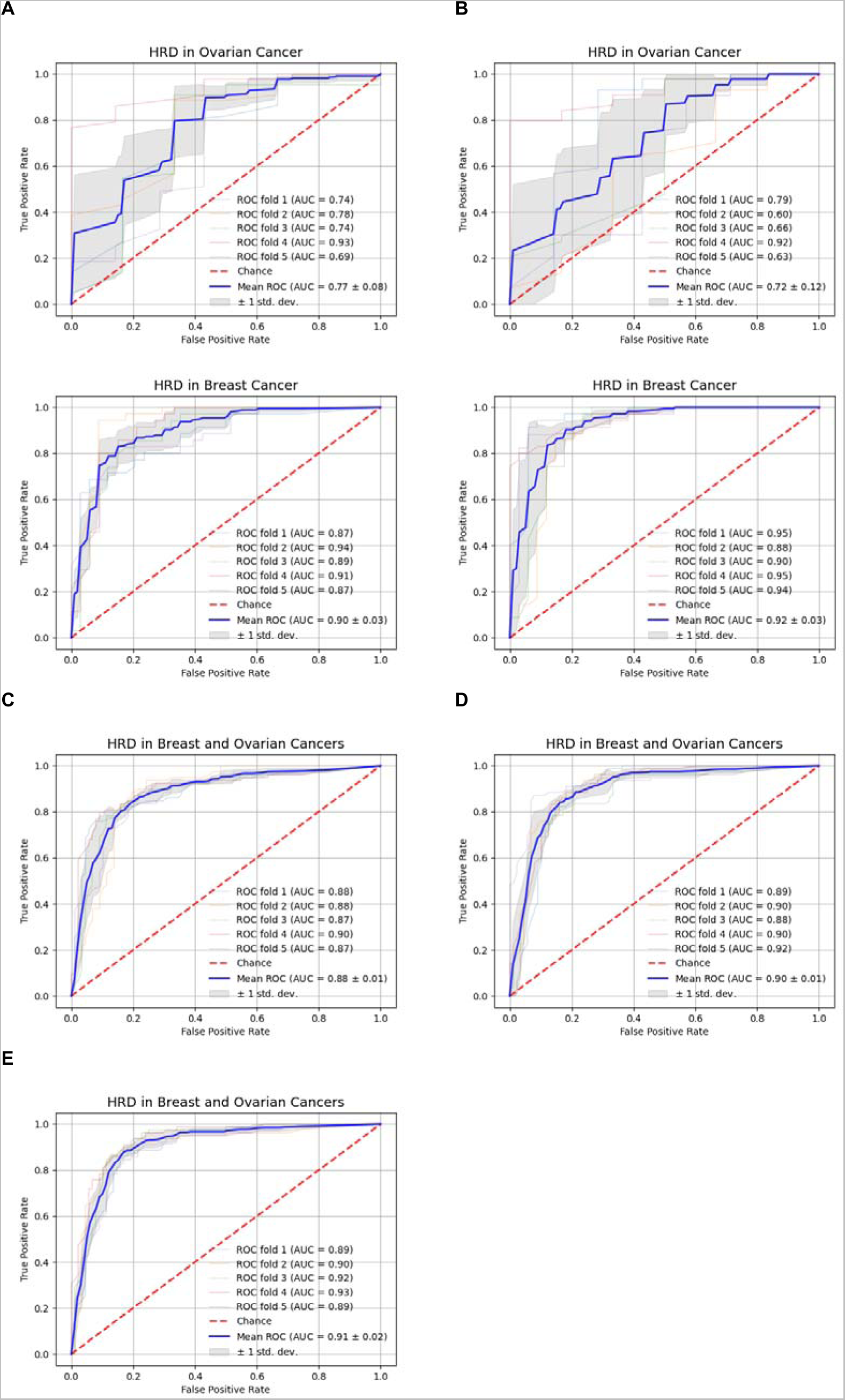
Classification models to predict homologous recombination deficiency using nullomers. Homologous recombination deficiency detection for: **A.** 14bp nullomers, **B.** 15bp nullomers. Homologous recombination for both breast and ovarian cancers together for **C.** 14bp nullomers, **D.** 15bp nullomers, **E.** 16bp nullomers.

**Supplementary Figure 11:**
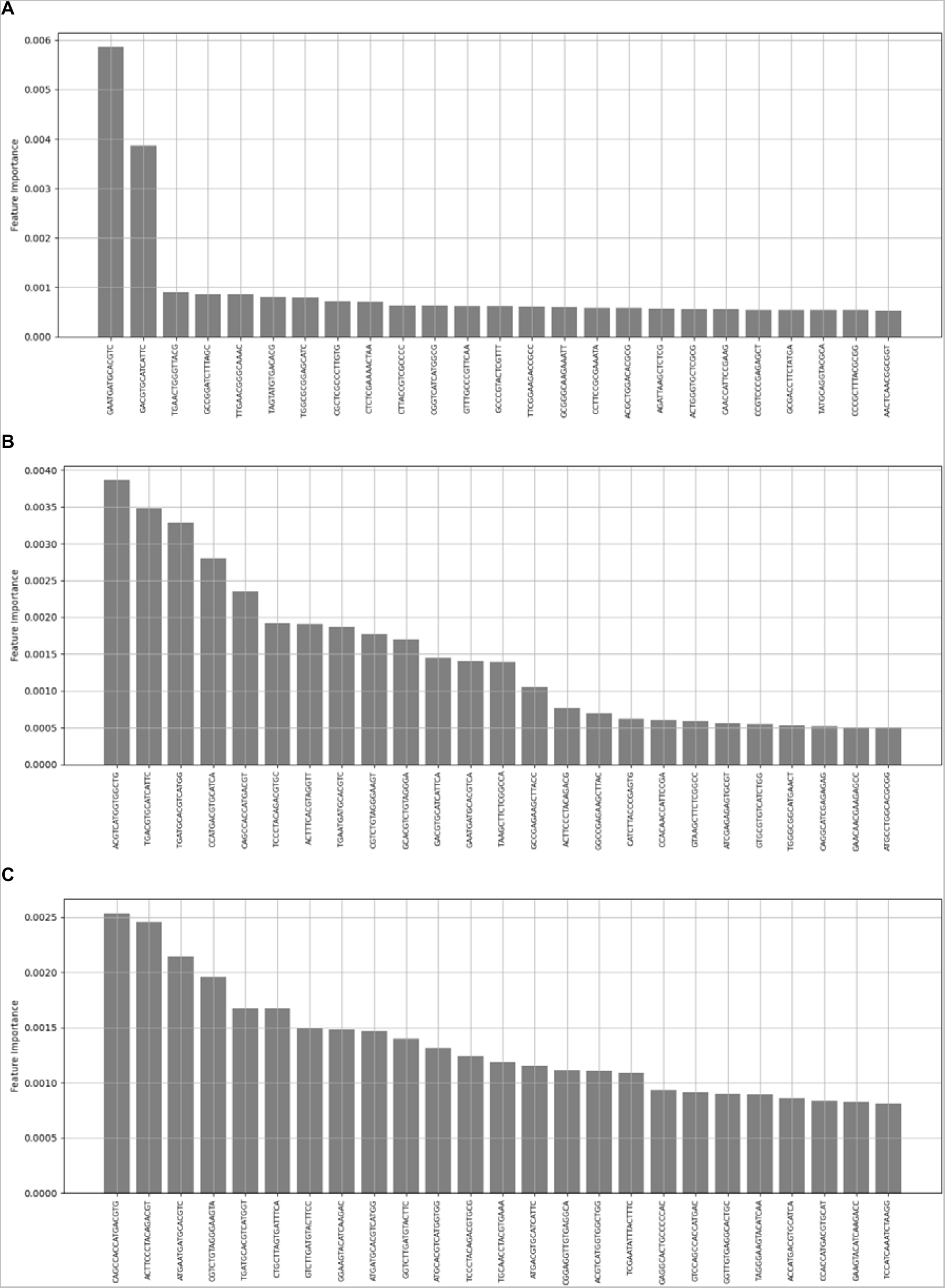
Homologous recombination deficiency detection feature importance from the classification model for: **A.** 14bp nullomers, **B.** 15bp nullomers, **C.** 16bp nullomers.

**Supplementary Figure 12:**
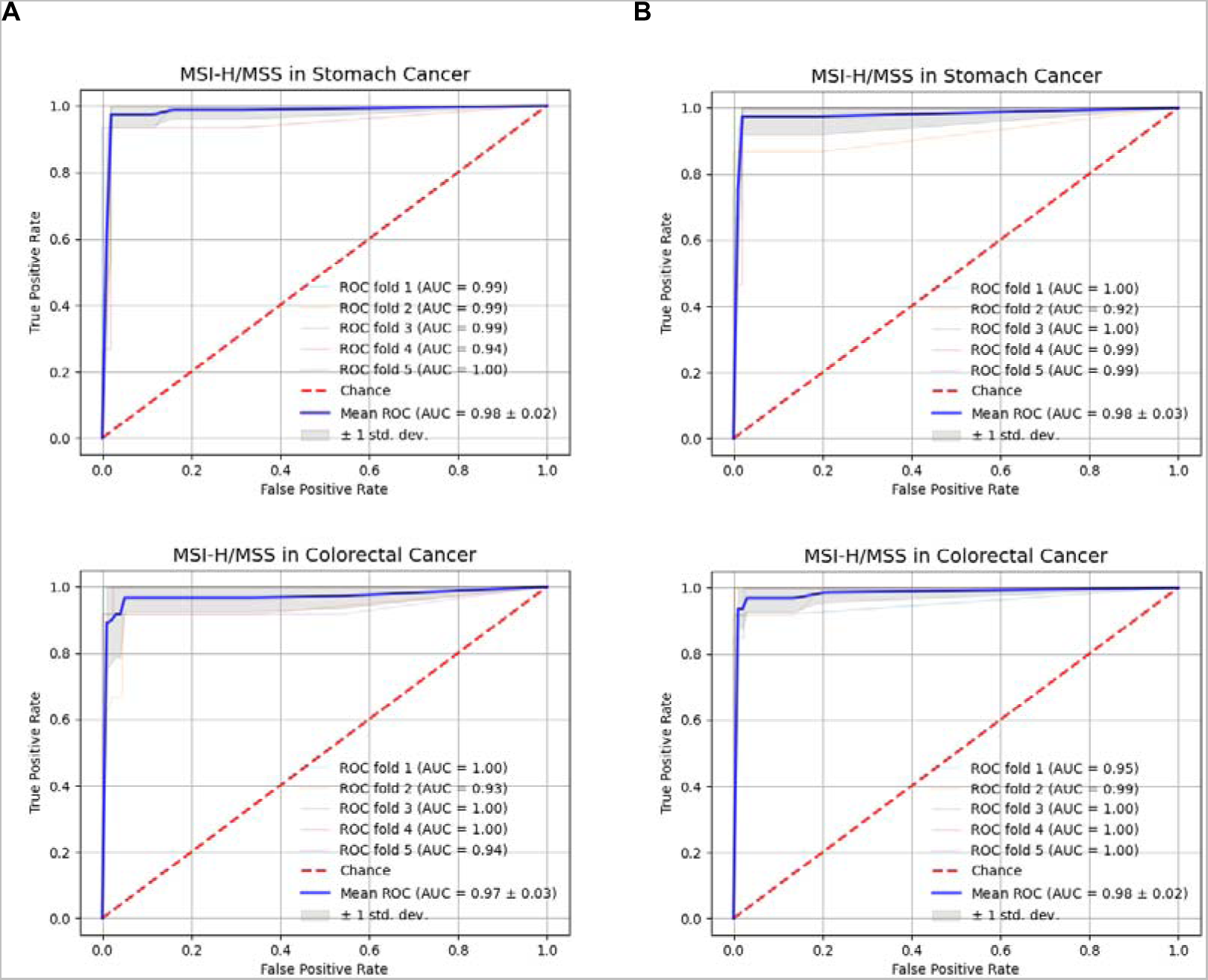
Mismatch repair deficiency detection using: **A.** 14bp nullomers, **B.** 15bp nullomers in stomach and colorectal cancers.

**Supplementary Figure 13:**
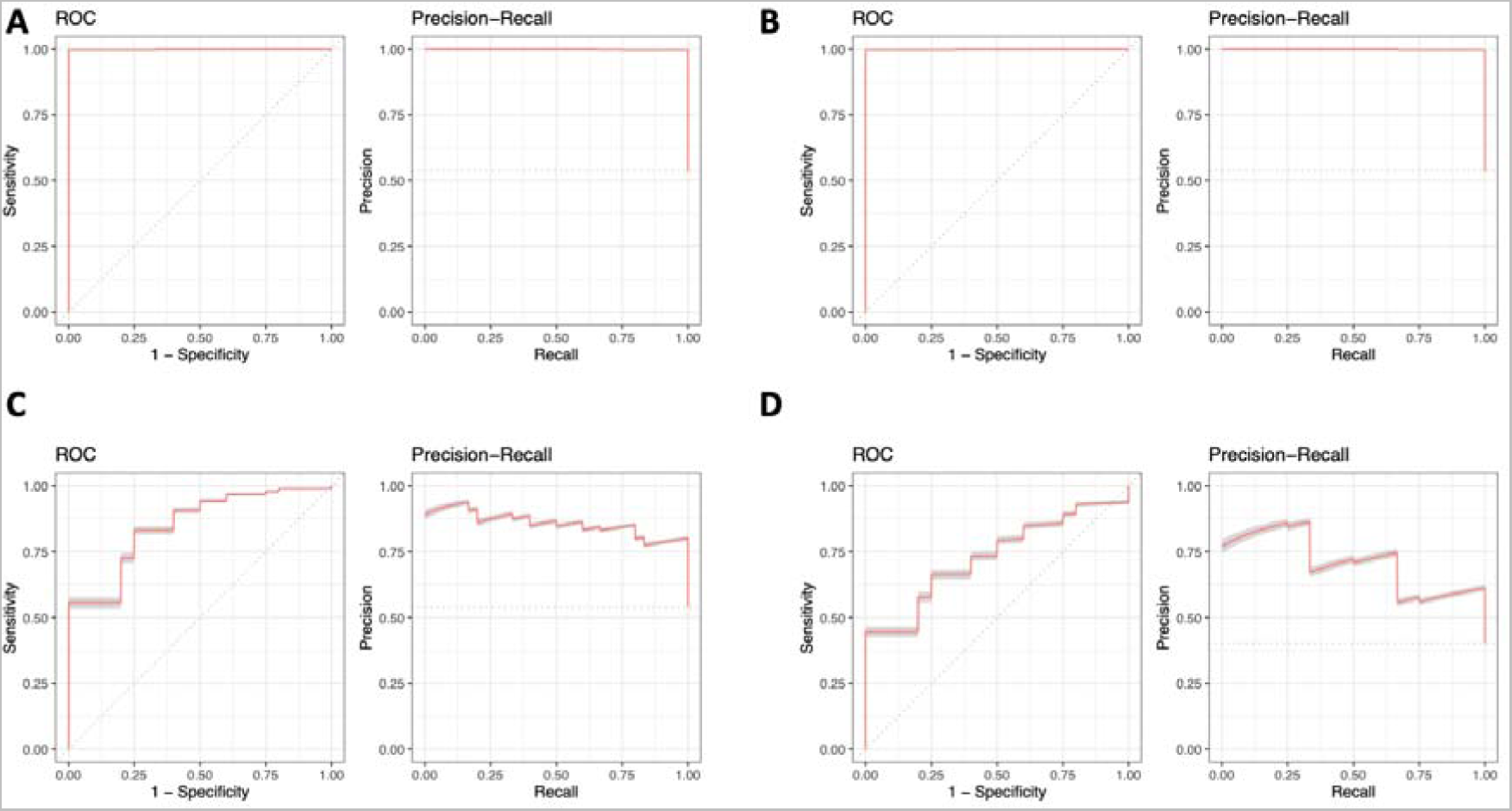
Predictive accuracy of cancer detection across cancer types. ROC curve and precision recall for: **A.** HCC for 14mers, **B.** HCC for 15mers, **C.** colorectal cancer, **D.** esophageal cancer

**Supplementary Figure 14:**
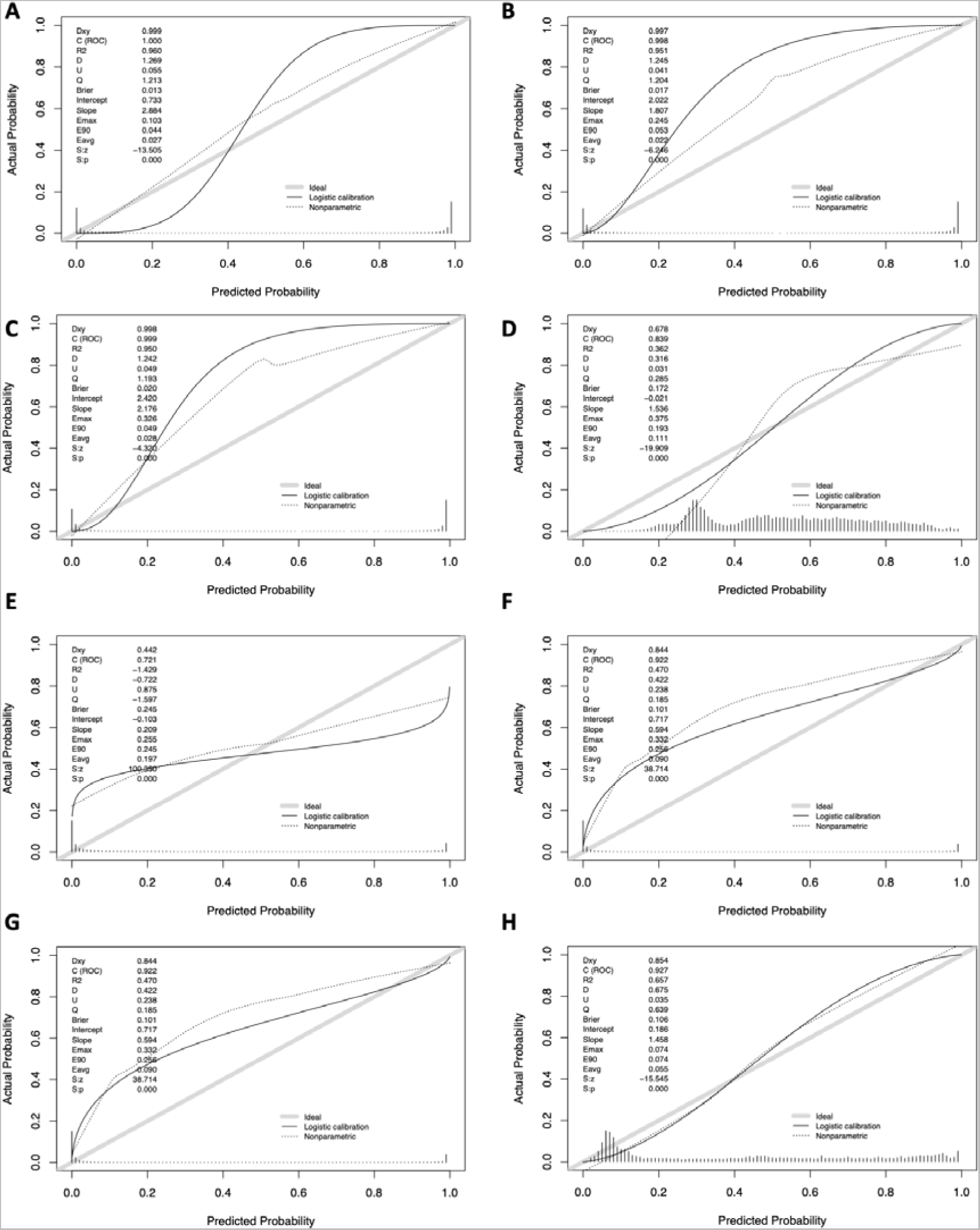
Calibration analysis to estimate performance of the cancer detection models. Results shown for HCC cancer **A.-C.** using sixteen, fifteen-mer and fourteen-mer nullomers and **D.-H.** using sixteen-mer nullomers for **D.** colorectal, **E.** esophageal, **F.** lung, **G.** liver and **H.** stomach cancers.

**Supplementary Table 1:**
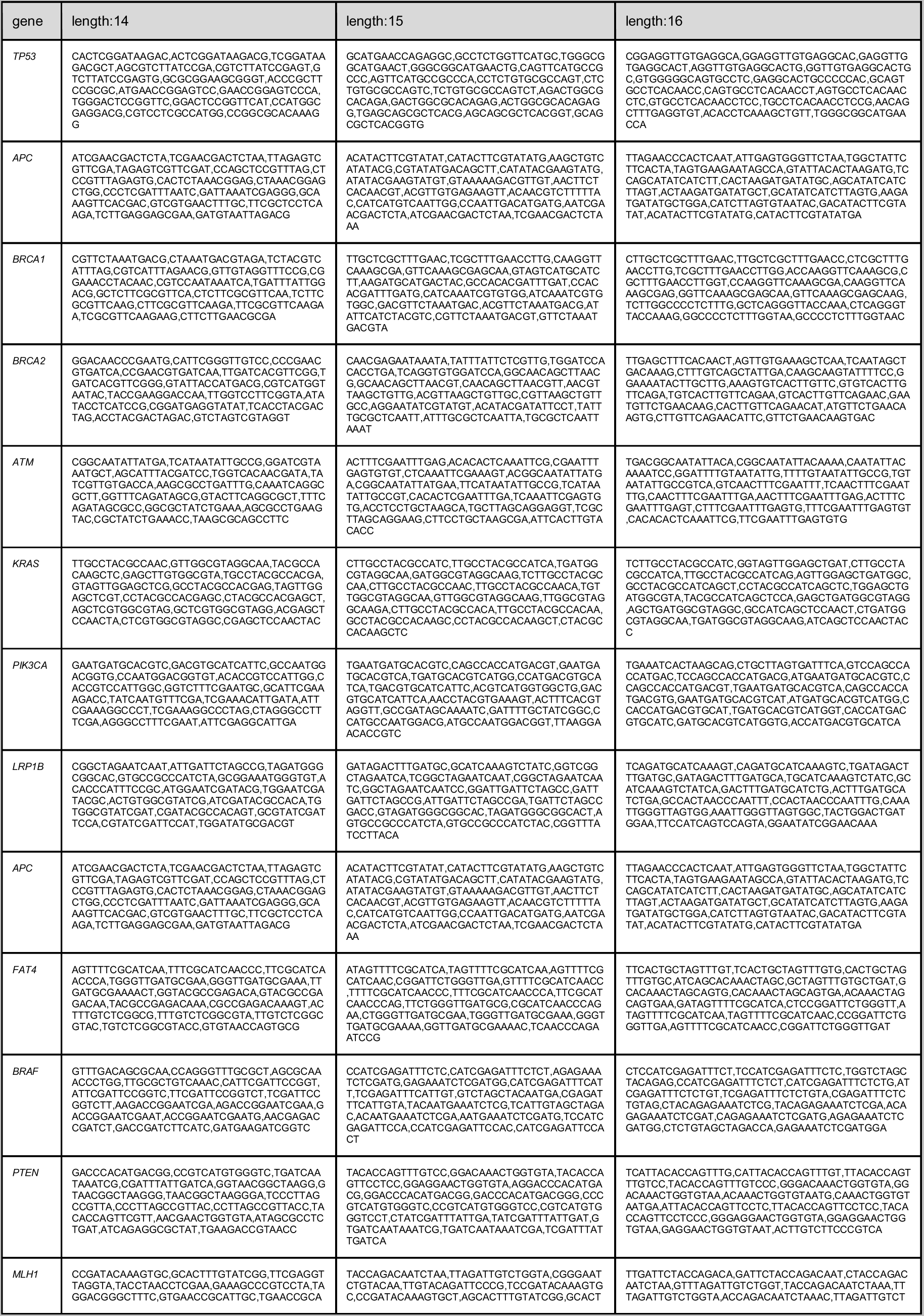
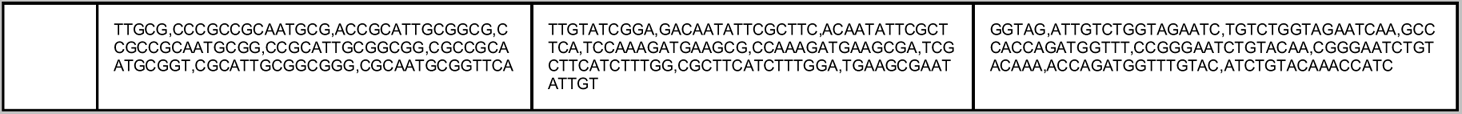
Up to fifteen highly frequent nullomers detected in selected top cancer genes for different kmer lengths.

**Supplementary Table 2:**
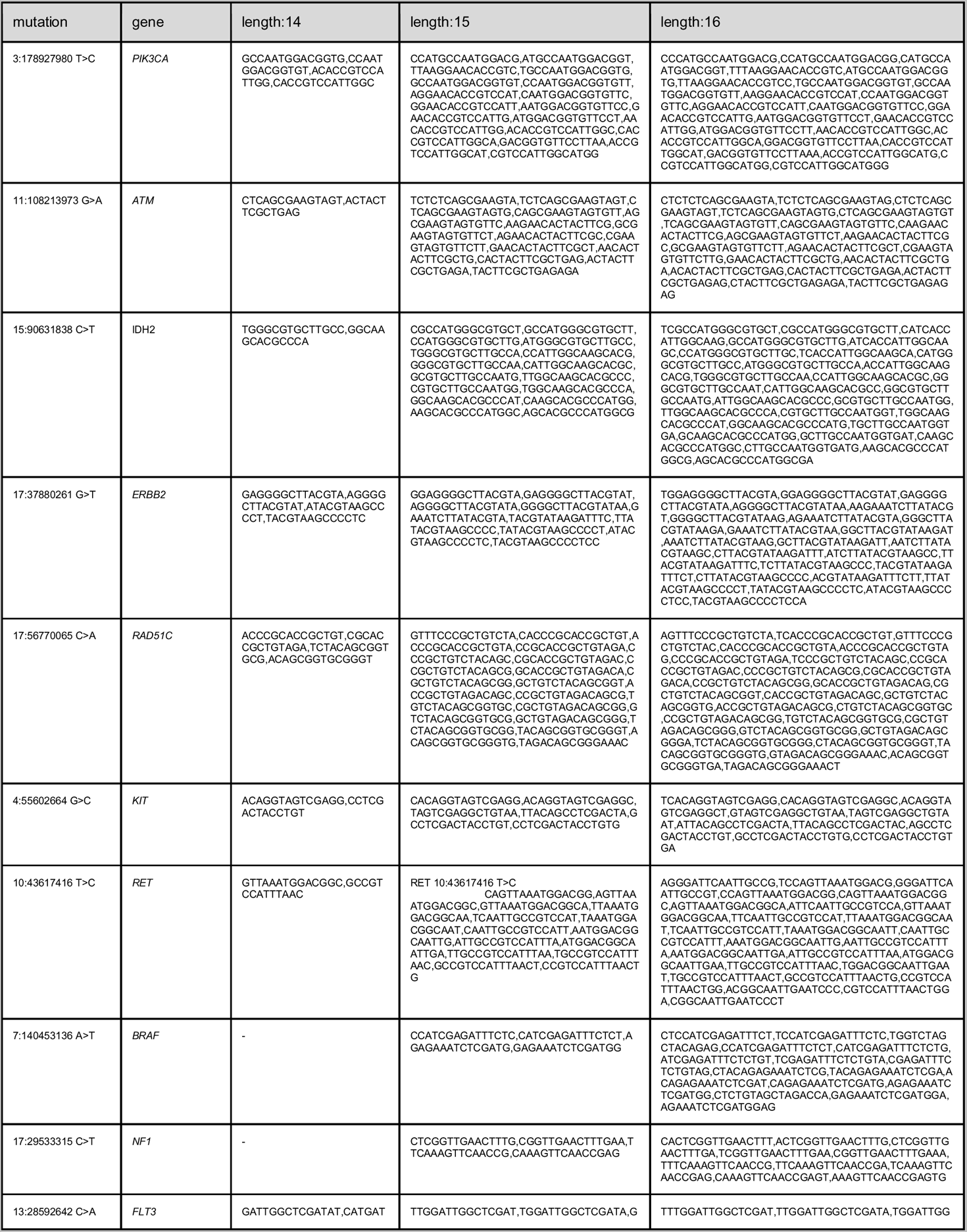
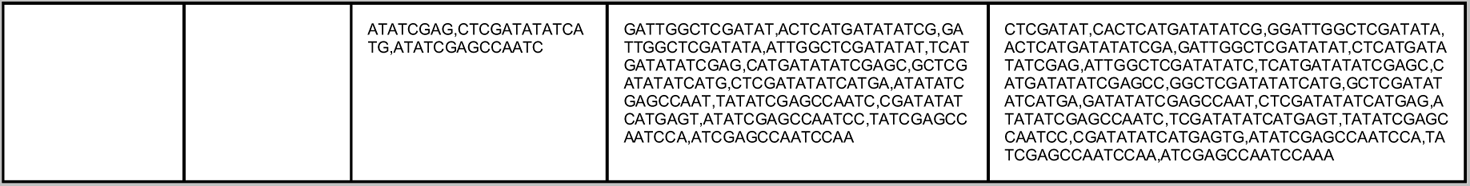
Selected clinically actionable mutations (classified as level 1, with FDA therapeutic drugs) and the nullomers that resurface from OncoKB (coordinates shown in hg19 reference genome).

**Supplementary Table 3:**
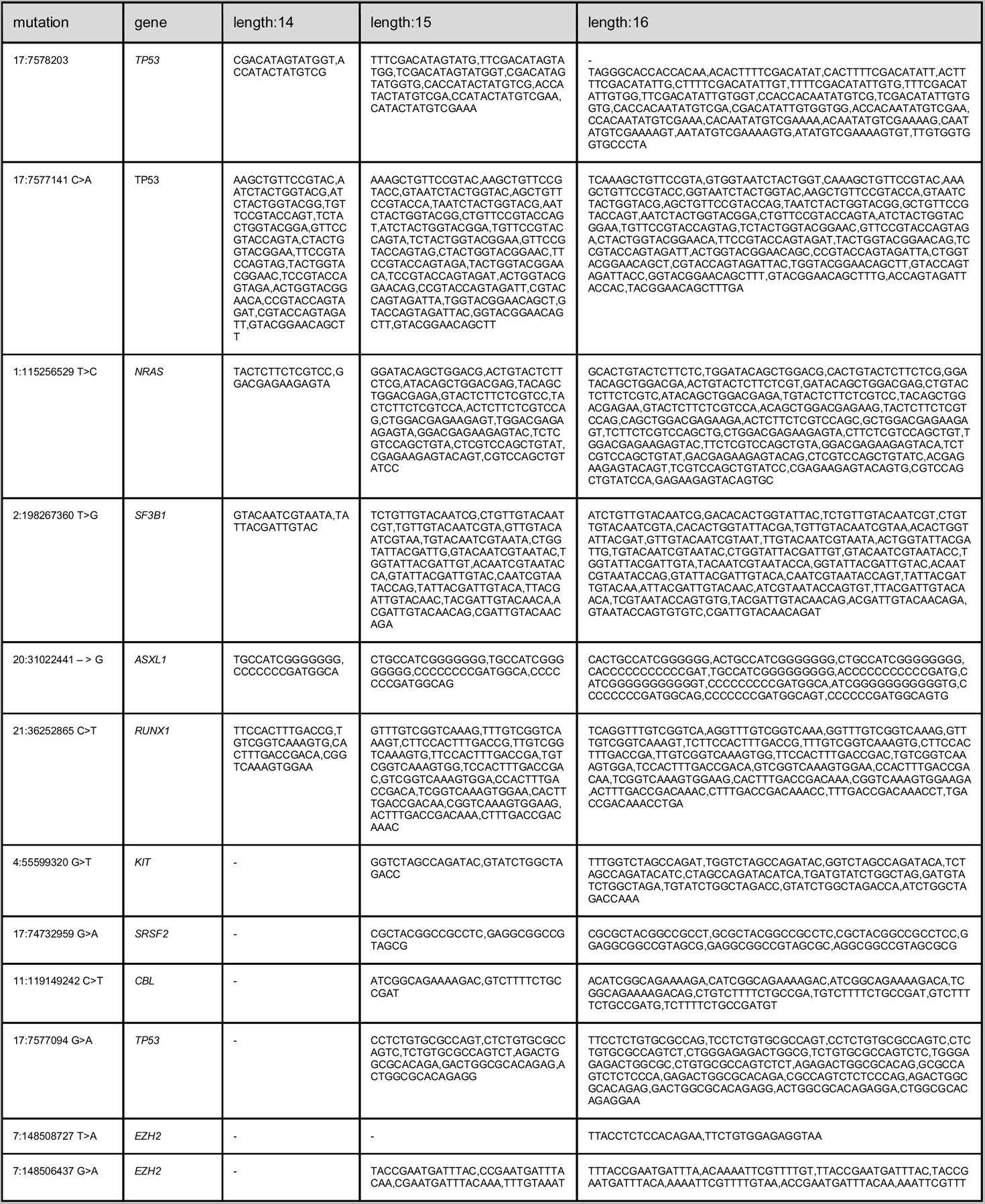
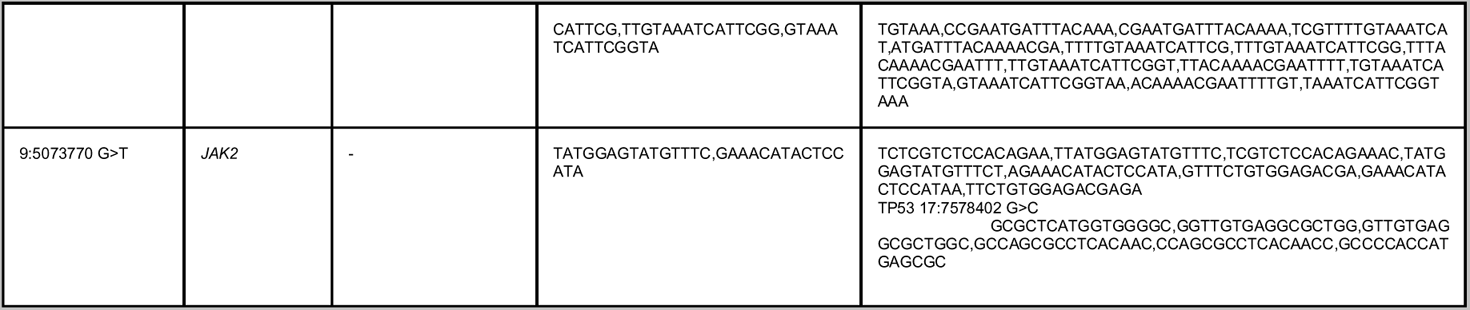
Selected clinically actionable mutations (classified as level 1, for prognosis) and the nullomers that resurface from OncoKB (coordinates shown in hg19 reference genome).

**Supplementary Table 4:**
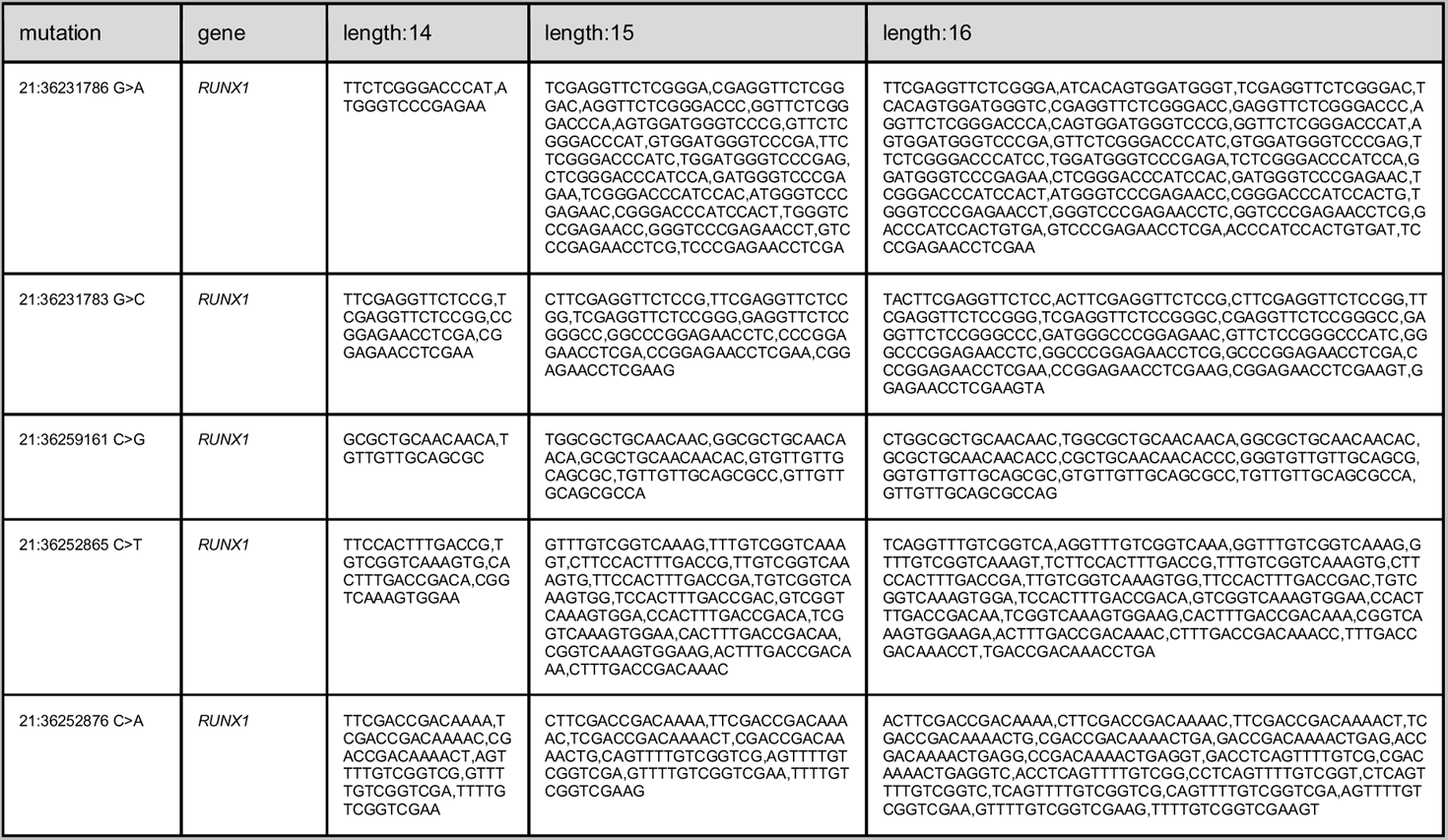
Selected clinically actionable mutations (classified as level 1, for diagnosis) and the nullomers that resurface from OncoKB (coordinates shown in hg19 reference genome).

**Supplementary Table 5:**
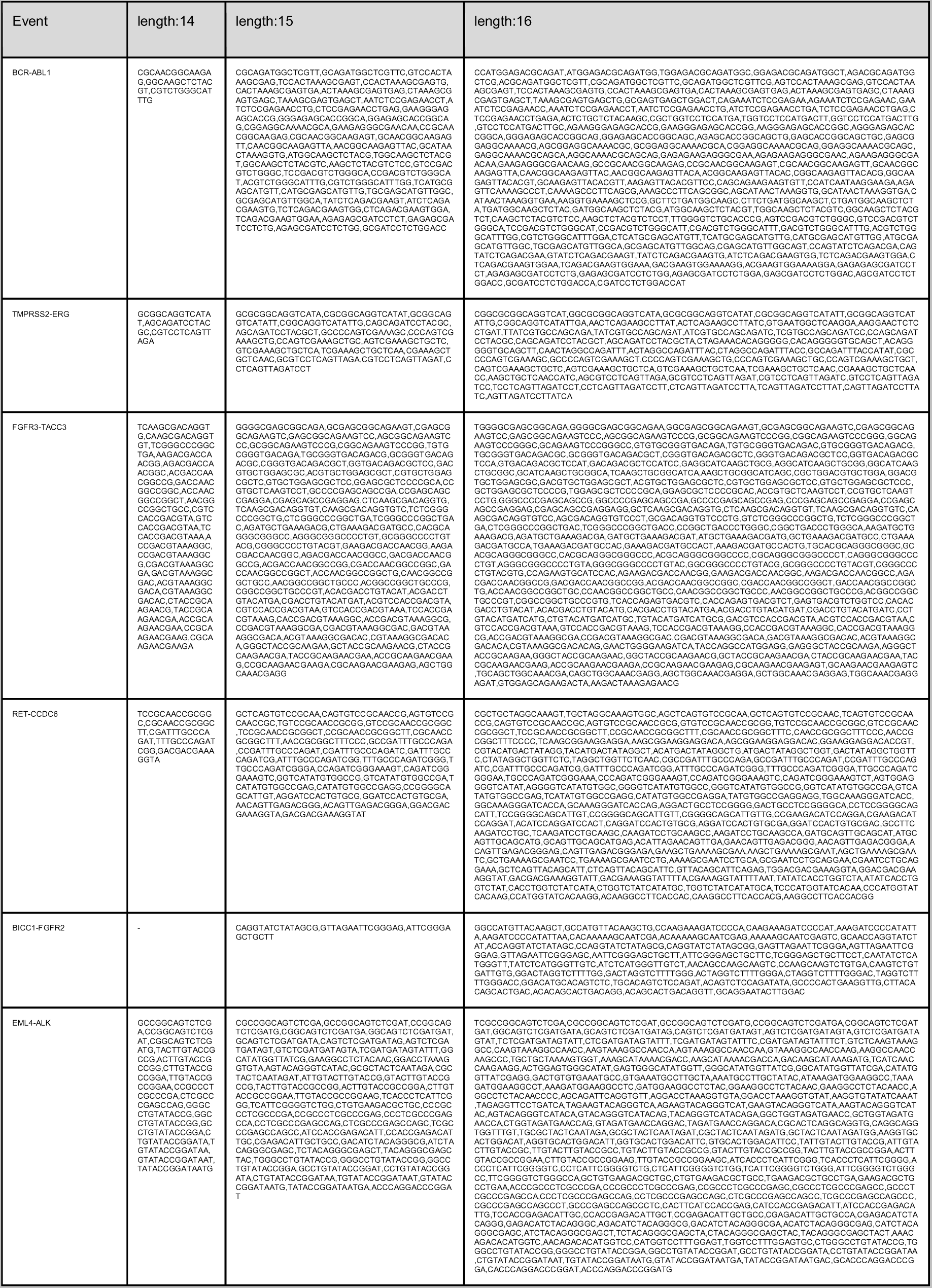
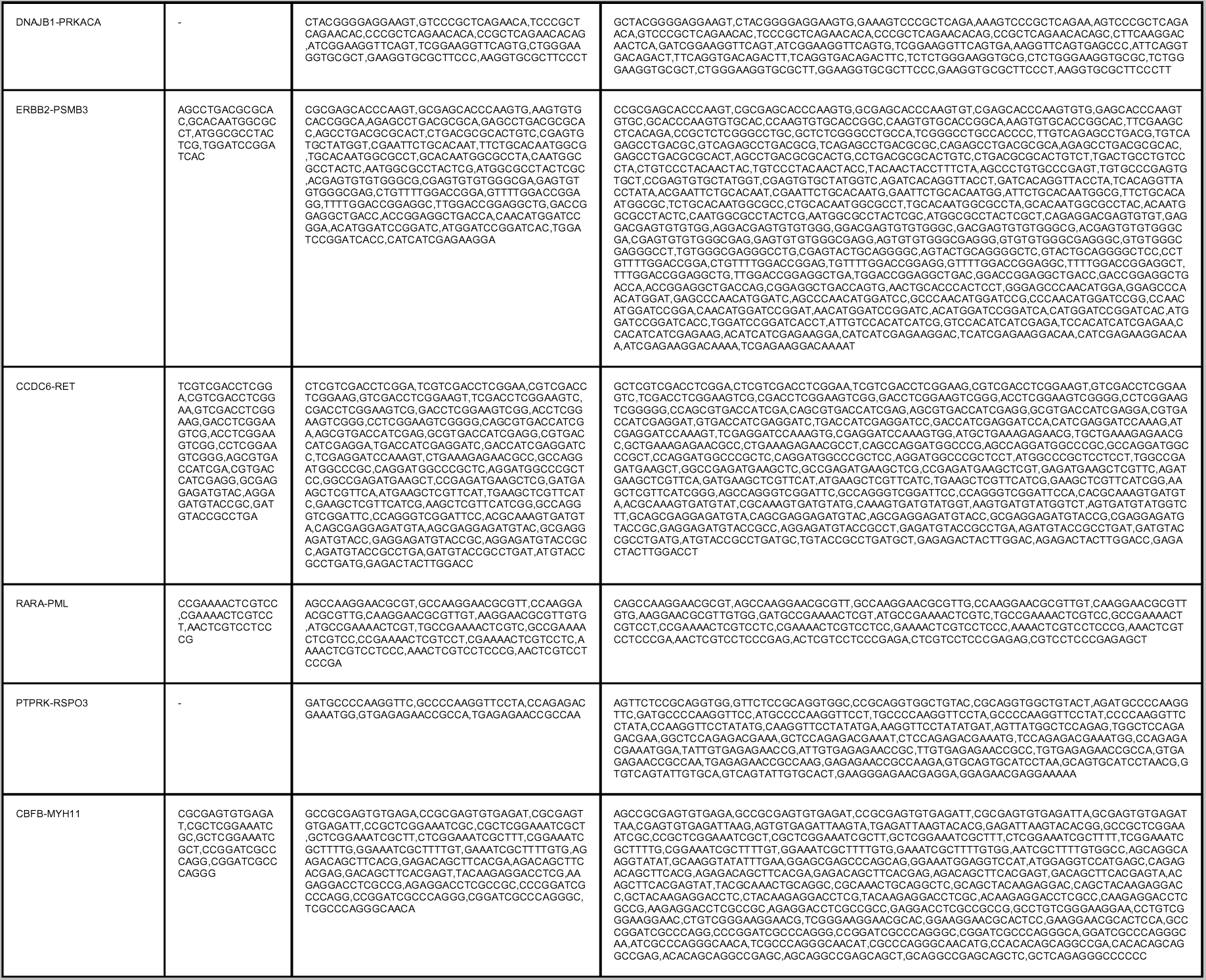
Nullomer resurfacing from fusion events.

**Supplementary Table 6:**
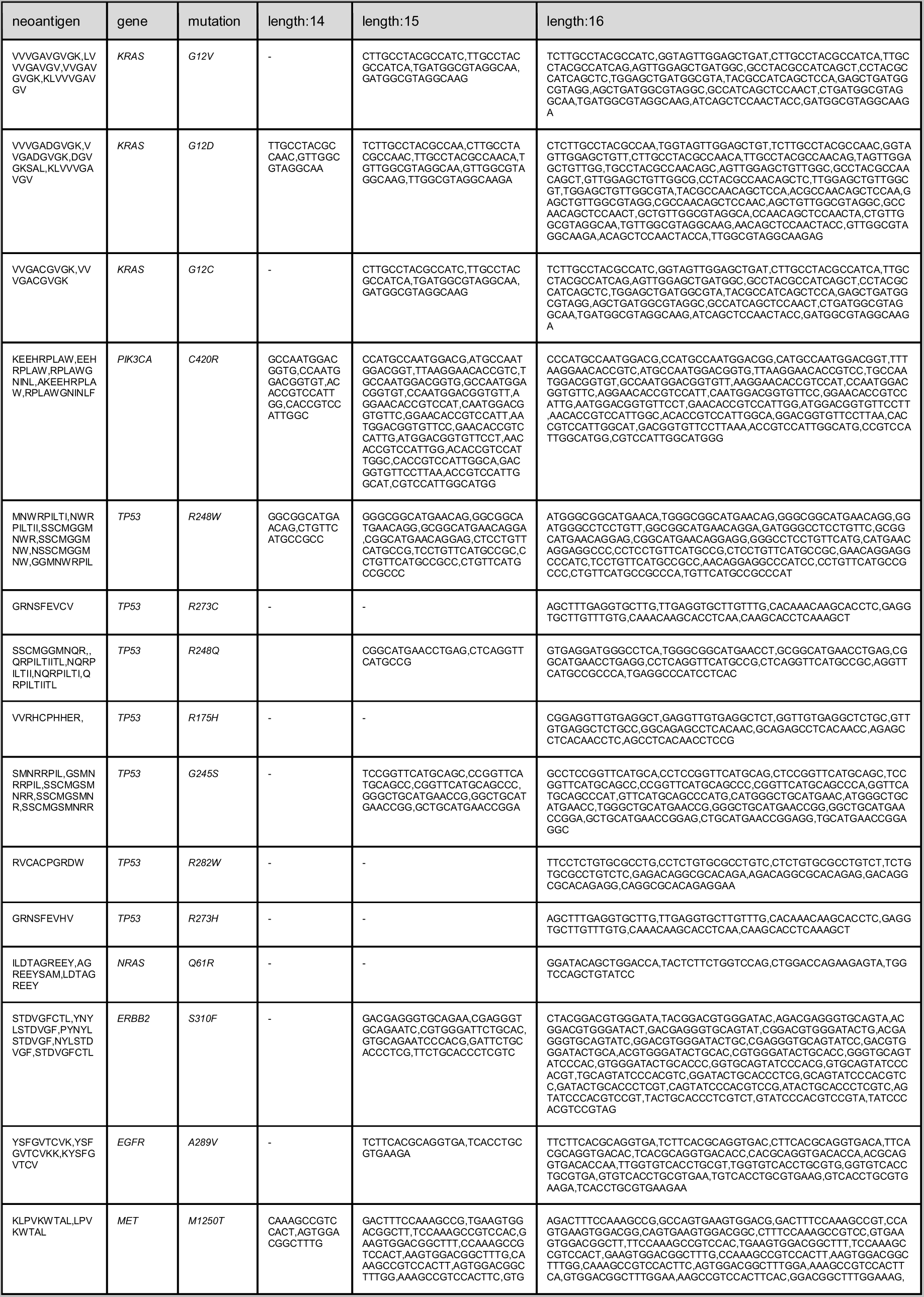
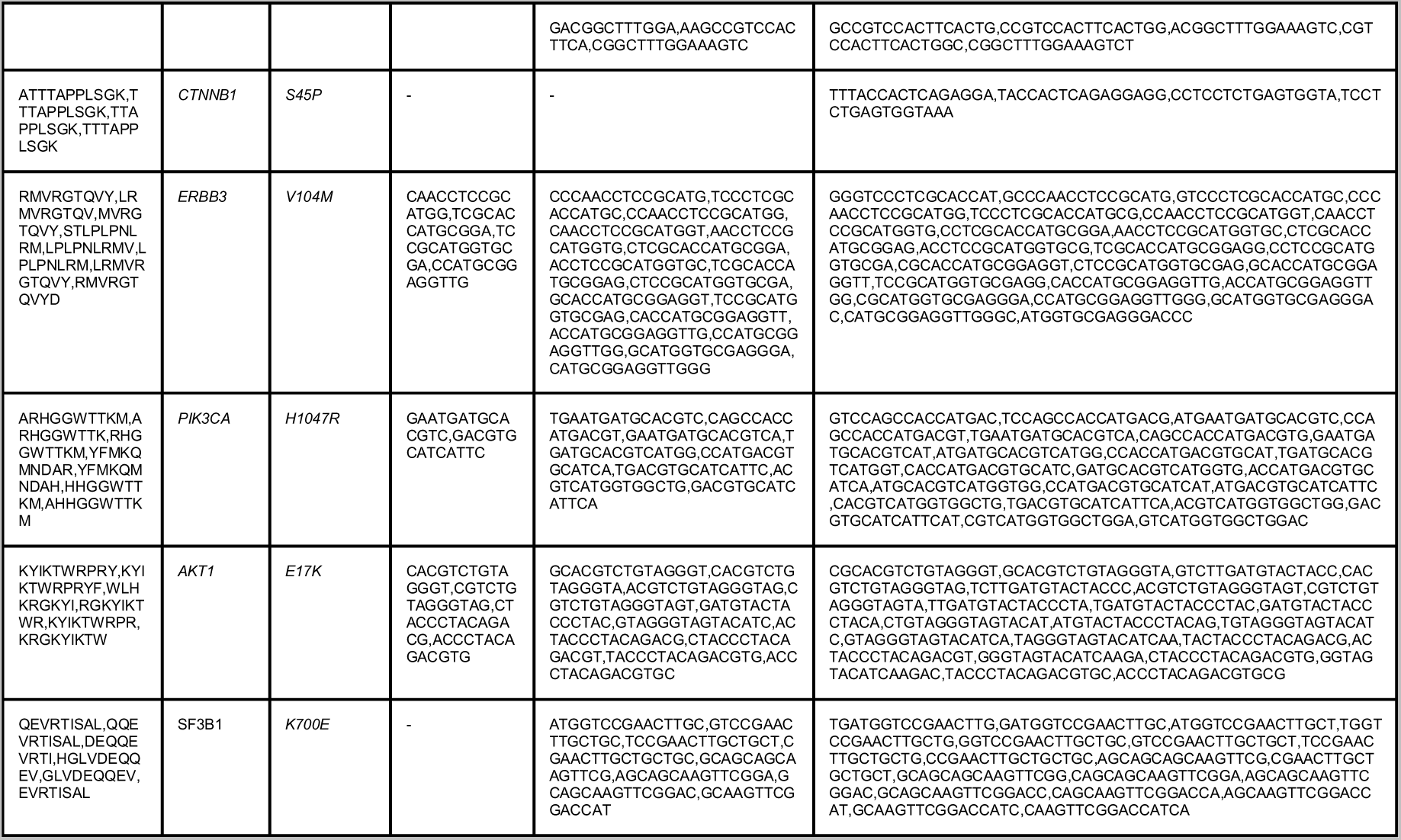
Selected neoantigens and the nullomers that resurface from the mutations that generate them.

## Code availability

The code for this work can be found at: https://github.com/Georgakopoulos-Soares-lab/cfRNA-nullomer-analysis

## Contributions

I.G.S. conceived the study. A.M., G.C.T, I.M., M.A., and I.G.S. wrote the code. A.M., G.C.T., and I.G.S. generated the schematics. A.M., G.C.T., I.M., and I.G.S. wrote the manuscript with inputs from all the authors.

## Acknowledgements

This study was funded by the startup funds of I.G.S. from the Penn State College of Medicine.

## Bibliography

Acquisti, Claudia, George Poste, David Curtiss, and Sudhir Kumar. 2007. “Nullomers: Really a Matter of Natural Selection?” PloS One 2 (10): e1022.

Bushnell, Brian, Jonathan Rood, and Esther Singer. 2017. “BBMerge - Accurate Paired Shotgun Read Merging via Overlap.” PloS One 12 (10): e0185056.

Chakravarty, Debyani, Jianjiong Gao, Sarah M. Phillips, Ritika Kundra, Hongxin Zhang, Jiaojiao Wang, Julia E. Rudolph, et al. 2017. “OncoKB: A Precision Oncology Knowledge Base.” JCO Precision Oncology 2017 (July). https://doi.org/10.1200/PO.17.00011.

Chen, Shanwen, Yunfan Jin, Siqi Wang, Shaozhen Xing, Yingchao Wu, Yuhuan Tao, Yongchen Ma, et al. 2022. “Cancer Type Classification Using Plasma Cell-Free RNAs Derived from Human and Microbes.” eLife 11 (July). https://doi.org/10.7554/eLife.75181.

Chen, Shi-Lu, Shi-Xun Lu, Li-Li Liu, Chun-Hua Wang, Xia Yang, Zhi-Yi Zhang, Hui-Zhong Zhang, and Jing-Ping Yun. 2018. “eEF1A1 Overexpression Enhances Tumor Progression and Indicates Poor Prognosis in Hepatocellular Carcinoma.” Translational Oncology 11 (1): 125–31.

Crosby, David, Sangeeta Bhatia, Kevin M. Brindle, Lisa M. Coussens, Caroline Dive, Mark Emberton, Sadik Esener, et al. 2022. “Early Detection of Cancer.” Science 375 (6586): eaay9040.

Ding, Zhiyong, Nan Wang, Ning Ji, and Zhe-Sheng Chen. 2022. “Proteomics Technologies for Cancer Liquid Biopsies.” Molecular Cancer 21 (1): 53.

Ellrott, Kyle, Matthew H. Bailey, Gordon Saksena, Kyle R. Covington, Cyriac Kandoth, Chip Stewart, Julian Hess, et al. 2018. “Scalable Open Science Approach for Mutation Calling of Tumor Exomes Using Multiple Genomic Pipelines.” Cell Systems 6 (3): 271–81.e7.

Enderle, Daniel, Alexandra Spiel, Christine M. Coticchia, Emily Berghoff, Romy Mueller, Martin Schlumpberger, Markus Sprenger-Haussels, et al. 2015. “Characterization of RNA from Exosomes and Other Extracellular Vesicles Isolated by a Novel Spin Column-Based Method.” PloS One 10 (8): e0136133.

Friedman, Jerome, Trevor Hastie, and Rob Tibshirani. 2010. “Regularization Paths for Generalized Linear Models via Coordinate Descent.” Journal of Statistical Software 33 (1): 1–22.

Georgakopoulos-Soares, Ilias, Ofer Yizhar Barnea, Ioannis Mouratidis, Rachael Bradley, Ryder Easterlin, Candace Chan, Emmalyn Chen, John S. Witte, Martin Hemberg, and Nadav Ahituv. 2021. “Leveraging Sequences Missing from the Human Genome to Diagnose Cancer.” medRxiv.

Georgakopoulos-Soares, Ilias, Ofer Yizhar-Barnea, Ioannis Mouratidis, Martin Hemberg, and Nadav Ahituv. 2021. “Absent from DNA and Protein: Genomic Characterization of Nullomers and Nullpeptides across Functional Categories and Evolution.” Genome Biology 22 (1): 245.

Hou, Guoxin, Zhimin Lu, Jialu Jiang, and Xinmei Yang. 2023. “Ribosomal Protein L32 Enhances Hepatocellular Carcinoma Progression.” Cancer Medicine, April. https://doi.org/10.1002/cam4.5811.

Hu, Wanye, Chaoting Zhou, Qiangan Jing, Yancun Li, Jing Yang, Chen Yang, Luyang Wang, et al. 2021. “FTH Promotes the Proliferation and Renders the HCC Cells Specifically Resist to Ferroptosis by Maintaining Iron Homeostasis.” Cancer Cell International 21 (1): 709.

Ignatiadis, Michail, George W. Sledge, and Stefanie S. Jeffrey. 2021. “Liquid Biopsy Enters the Clinic - Implementation Issues and Future Challenges.” Nature Reviews. Clinical Oncology 18 (5): 297–312.

Islam, S. M. Ashiqul, Marcos Díaz-Gay, Yang Wu, Mark Barnes, Raviteja Vangara, Erik N. Bergstrom, Yudou He, et al. 2022. “Uncovering Novel Mutational Signatures by Extraction with SigProfilerExtractor.” Cell Genomics 2 (11): None.

Jang, Ye Eun, Insu Jang, Sunkyu Kim, Subin Cho, Daehan Kim, Keonwoo Kim, Jaewon Kim, et al. 2020. “ChimerDB 4.0: An Updated and Expanded Database of Fusion Genes.” Nucleic Acids Research 48 (D1): D817–24.

Knight, John Rp, Nikola Vlahov, David M. Gay, Rachel A. Ridgway, William James Faller, Christopher Proud, Giovanna R. Mallucci, et al. 2021. “Mutation Suppresses Colorectal Cancer by Promoting eEF2 Phosphorylation via eEF2K.” eLife 10 (December). https://doi.org/10.7554/eLife.69729.

Konstantinopoulos, Panagiotis A., Raphael Ceccaldi, Geoffrey I. Shapiro, and Alan D. D’Andrea. 2015. “Homologous Recombination Deficiency: Exploiting the Fundamental Vulnerability of Ovarian Cancer.” Cancer Discovery 5 (11): 1137–54.

Koulouras, Grigorios, and Martin C. Frith. 2021. “Significant Non-Existence of Sequences in Genomes and Proteomes.” Nucleic Acids Research 49 (6): 3139–55.

Krueger, Felix, Frankie James, Phil Ewels, Ebrahim Afyounian, Michael Weinstein, Benjamin Schuster-Boeckler, Gert Hulselmans, and sclamons. 2023. FelixKrueger/TrimGalore: v0.6.10 - Add Default Decompression Path. Zenodo. https://doi.org/10.5281/ZENODO.7598955.

Kuhn, Max. 2008. “Building Predictive Models in R Using the Caret Package.” Journal of Statistical Software 28 (November): 1–26.

Lang, Franziska, Barbara Schrörs, Martin Löwer, Özlem Türeci, and Ugur Sahin. 2022. “Identification of Neoantigens for Individualized Therapeutic Cancer Vaccines.” Nature Reviews. Drug Discovery 21 (4): 261–82.

Larson, Matthew H., Wenying Pan, Hyunsung John Kim, Ruth E. Mauntz, Sarah M. Stuart, Monica Pimentel, Yiqi Zhou, et al. 2021. “A Comprehensive Characterization of the Cell-Free Transcriptome Reveals Tissue- and Subtype-Specific Biomarkers for Cancer Detection.” Nature Communications 12 (1): 2357.

Larson, Matthew H., Wenying Pan, Hyunsung John Kim, Ruth E. Mauntz, Sarah M. Stuart, Monica Pimentel, Yiqi Zhou, et al. 2022. “Author Correction: A Comprehensive Characterization of the Cell-Free Transcriptome Reveals Tissue- and Subtype-Specific Biomarkers for Cancer Detection.” Nature Communications 13 (1): 2553.

Li, Guangyao, Sehar Samuel, Sher Ehsan Ul Haq, Ayman S. Mubarak, Christian R. Studenik, Asif Islam, Mohammed Aufy, Mustafa Jawad Kadham, Abdul Kareem J. Al-Azzawi, and Mostafa A. Abdel-Maksoud. 2023. “Characterizing the Oncogenic Importance and Exploring Gene-Immune Cells Correlation of ACTB in Human Cancers.” American Journal of Cancer Research 13 (3): 758–77.

Liu, Yang, Nilay S. Sethi, Toshinori Hinoue, Barbara G. Schneider, Andrew D. Cherniack, Francisco Sanchez-Vega, Jose A. Seoane, et al. 2018. “Comparative Molecular Analysis of Gastrointestinal Adenocarcinomas.” Cancer Cell 33 (4): 721–35.e8.

Locke, Warwick J., Dominic Guanzon, Chenkai Ma, Yi Jin Liew, Konsta R. Duesing, Kim Y. C. Fung, and Jason P. Ross. 2019. “DNA Methylation Cancer Biomarkers: Translation to the Clinic.” Frontiers in Genetics 10 (November): 1150.

Lone, Saife N., Sabah Nisar, Tariq Masoodi, Mayank Singh, Arshi Rizwan, Sheema Hashem, Wael El-Rifai, et al. 2022. “Liquid Biopsy: A Step Closer to Transform Diagnosis, Prognosis and Future of Cancer Treatments.” Molecular Cancer 21 (1): 79.

Marçais, Guillaume, and Carl Kingsford. 2011. “A Fast, Lock-Free Approach for Efficient Parallel Counting of Occurrences of K-Mers.” Bioinformatics 27 (6): 764–70.

Mattiuzzi, Camilla, and Giuseppe Lippi. 2019. “Current Cancer Epidemiology.” Journal of Epidemiology and Global Health 9 (4): 217–22.

Mertens, Fredrik, Bertil Johansson, Thoas Fioretos, and Felix Mitelman. 2015. “The Emerging Complexity of Gene Fusions in Cancer.” Nature Reviews. Cancer 15 (6): 371–81.

Nielsen, Jens. 2017. “Systems Biology of Metabolism: A Driver for Developing Personalized and Precision Medicine.” Cell Metabolism 25 (3): 572–79.

Pan, Biran, Tongtong Zhang, Wei Yang, Yanjun Liu, Yuning Chen, Zheng Zhou, Yan Tang, et al. 2019. “SNX3 Suppresses the Migration and Invasion of Colorectal Cancer Cells by Reversing Epithelial-to-Mesenchymal Transition via the β-Catenin Pathway.” Oncology Letters 18 (5): 5332–40.

Peng, Mingyu, Lin Ye, Li Yang, Xinzhu Liu, Yuhan Chen, Guichuan Huang, Yu Jiang, et al. 2020. “Is Frequently Silenced by CpG Methylation and Sensitizes Lung Cancer Cells to Paclitaxel and 5-FU.” Epigenomics 12 (20): 1793–1810.

Peng, Ningfu, Jingrong He, Jindu Li, Hao Huang, Weiqiao Huang, Yingyang Liao, and Shaoliang Zhu. 2020. “Long Noncoding RNA MALAT1 Inhibits the Apoptosis and Autophagy of Hepatocellular Carcinoma Cell by Targeting the microRNA-146a/PI3K/Akt/mTOR Axis.” Cancer Cell International 20 (May): 165.

Robinson, Mark D., Davis J. McCarthy, and Gordon K. Smyth. 2010. “edgeR: A Bioconductor Package for Differential Expression Analysis of Digital Gene Expression Data.” Bioinformatics 26 (1): 139–40.

Ruiz-Iglesias, Ainhoa, and Santos Mañes. 2021. “The Importance of Mitochondrial Pyruvate Carrier in Cancer Cell Metabolism and Tumorigenesis.” Cancers 13 (7). https://doi.org/10.3390/cancers13071488.

Rushton, Amelia J., Georgios Nteliopoulos, Jacqueline A. Shaw, and R. Charles Coombes. 2021. “A Review of Circulating Tumour Cell Enrichment Technologies.” Cancers 13 (5). https://doi.org/10.3390/cancers13050970.

Saito, Takaya, and Marc Rehmsmeier. 2017. “Precrec: Fast and Accurate Precision-Recall and ROC Curve Calculations in R.” Bioinformatics 33 (1): 145–47.

Scarpa, Emanuele Salvatore, Filippo Tasini, Rita Crinelli, Chiara Ceccarini, Mauro Magnani, and Marzia Bianchi. 2020. “The Ubiquitin Gene Expression Pattern and Sensitivity to and Knockdown Differentiate Primary 23132/87 and Metastatic MKN45 Gastric Cancer Cells.” International Journal of Molecular Sciences 21 (15). https://doi.org/10.3390/ijms21155435.

Siegel, Rebecca L., Kimberly D. Miller, and Ahmedin Jemal. 2020. “Cancer Statistics, 2020.” CA: A Cancer Journal for Clinicians 70 (1): 7–30.

Song, Chunrong, Zhong Su, and Jing Guo. 2019. “Thymosin β 10 Is Overexpressed and Associated with Unfavorable Prognosis in Hepatocellular Carcinoma.” Bioscience Reports 39 (3). https://doi.org/10.1042/BSR20182355.

Stratton, Michael R., Peter J. Campbell, and P. Andrew Futreal. 2009. “The Cancer Genome.” Nature 458 (7239): 719–24.

Telli, Melinda L., Kirsten M. Timms, Julia Reid, Bryan Hennessy, Gordon B. Mills, Kristin C. Jensen, Zoltan Szallasi, et al. 2016. “Homologous Recombination Deficiency (HRD) Score Predicts Response to Platinum-Containing Neoadjuvant Chemotherapy in Patients with Triple-Negative Breast Cancer.” Clinical Cancer Research: An Official Journal of the American Association for Cancer Research 22 (15): 3764–73.

Thorsson, Vésteinn, David L. Gibbs, Scott D. Brown, Denise Wolf, Dante S. Bortone, Tai-Hsien Ou Yang, Eduard Porta-Pardo, et al. 2018. “The Immune Landscape of Cancer.” Immunity 48 (4): 812–30.e14.

Whitehurst, Angelique W., Yang Xie, Scott C. Purinton, Kathryn M. Cappell, Jackie T. Swanik, Brittany Larson, Luc Girard, John O. Schorge, and Michael A. White. 2010. “Tumor Antigen Acrosin Binding Protein Normalizes Mitotic Spindle Function to Promote Cancer Cell Proliferation.” Cancer Research 70 (19): 7652– 61.

Wu, Jingcheng, Wenfan Chen, Yuxuan Zhou, Ying Chi, Xiansheng Hua, Jian Wu, Xun Gu, Shuqing Chen, and Zhan Zhou. 2022. “TSNAdb v2.0: The Updated Version of Tumor-Specific Neoantigen Database.” Genomics, Proteomics & Bioinformatics, October. https://doi.org/10.1016/j.gpb.2022.09.012.

Yao, Tian, Jin-Jin Liu, Li-Jun Zhao, Jing-Yi Zhou, Jia-Qi Wang, Yue Wang, Zhi-Qi Wang, Li-Hui Wei, Jian-Liu Wang, and Xiao-Ping Li. 2019. “Identification of New Fusion Genes and Their Clinical Significance in Endometrial Cancer.” Chinese Medical Journal 132 (11): 1314–21.

Yi, Ming, Dechao Jiao, Hanxiao Xu, Qian Liu, Weiheng Zhao, Xinwei Han, and Kongming Wu. 2018. “Biomarkers for Predicting Efficacy of PD-1/PD-L1 Inhibitors.” Molecular Cancer 17 (1): 129.

Zaporozhchenko, Ivan A., Anastasia A. Ponomaryova, Elena Yu Rykova, and Pavel P. Laktionov. 2018. “The Potential of Circulating Cell-Free RNA as a Cancer Biomarker: Challenges and Opportunities.” Expert Review of Molecular Diagnostics 18 (2): 133–45.

Zhao, Pengfei, Li Li, Xiaoyue Jiang, and Qin Li. 2019. “Mismatch Repair Deficiency/microsatellite Instability-High as a Predictor for Anti-PD-1/PD-L1 Immunotherapy Efficacy.” Journal of Hematology & Oncology 12 (1): 54.

Zhao, Peng, Fei Lan, Hui Zhang, Guangwei Zeng, and Dong Liu. 2018. “Down-Regulation of KIF2A Inhibits Gastric Cancer Cell Invasion via Suppressing MT1-MMP.” Clinical and Experimental Pharmacology & Physiology 45 (10): 1010–18.

Zhu, Yumin, Siqi Wang, Xiaochen Xi, Minfeng Zhang, Xiaofan Liu, Weina Tang, Peng Cai, et al. 2021. “Integrative Analysis of Long Extracellular RNAs Reveals a Detection Panel of Noncoding RNAs for Liver Cancer.” Theranostics 11 (1): 181–93.

